# COVID-19 in Children with Brain-Based Developmental Disabilities: A Rapid Review Update

**DOI:** 10.1101/2021.03.17.21253283

**Authors:** Michèle Dugas, Théo Stéfan, Johanie Lépine, Patrick Blouin, Andrée-Anne Poirier, Valérie Carnovale, Benoit Mailhot, Becky Skidmore, Lena Faust, Carrie Costello, Donna Thomson, Annette Majnemer, Dan Goldowitz, Steven P. Miller, Annie LeBlanc

## Abstract

**Objective:** Information regarding the impact of COVID-19 in children with brain-based disabilities, or those at risk of developing such conditions, remains scarce. The objective was to evaluate if children with brain-based disabilities are more likely to (1) develop COVID-19, (2) develop complications from the disease, and (3) to have a poorer prognosis.

**Study design:** We conducted a rapid review using search strategies iteratively developed and tested by an experienced medical information specialist in consultation with the review team and a panel of knowledge users. Searches were initially performed on April 18th, 2021, and updated on October 31st, 2020. Four reviewers individually performed study selection using pilot-tested standardized forms. Single reviewers extracted the data using a standardized extraction form that included study characteristics, patients’ characteristics, and outcomes reported.

**Results:** We identified 1448 publications, of which 29 were included. Studies reported data on 2288 COVID-19 positive children, including 462 with a brain-based disability, and 72 at risk of developing such disability. Overall, the included studies showed a greater risk to develop severe COVID-19 disease in children with brain-based disabilities. Although mortality is very low, the case-fatality rate appeared to be higher in children with disabilities compared to children without disabilities.

**Conclusions:** Our review shows that children with brain-based disabilities are overrepresented in hospitalization numbers compared to children without disabilities. However, most studies included children that were hospitalized from COVID-19 in secondary and tertiary care centers. Results of this review should therefore be interpreted with caution.

## Introduction

According to the World Health Organization, SARS-CoV-2 (COVID-19) has infected close to 105.4 million and caused the death of over 2.3 million individuals worldwide, as of February 7^th^, 2021^1^. Prevalence of COVID-19 in children is low. Of the 306,468 cases reported in Canada (November 17th, 2020), 42,527 (13.87%) were in patients aged 19 years old and under^2^. Of those, 273 were hospitalized (0.6%), and 47 were admitted to ICU (0.1%), with 2 deaths being reported. Data gathered for the U.S. indicated that of the 11,136,253 cases reported since the beginning of the pandemic, 787,558 (7.07%) were children, and of those, 117 death were reported^3^. Based on the current available evidence, children do not appear to be at higher risk of contracting COVID-19 than adults^4^. Symptom manifestation in children appears to be milder than in adults^5, 6^. Further, prevalence in individuals under 19 years of age has been quite low^5, 7, 8^ and children could even be less susceptible to the COVID-19 disease^4^. However, children with comorbidities may be vulnerable to severe COVID-19 disease^7^.

Certain individuals are at increased risk of experiencing severe outcomes or poorer prognoses, including older adults (aged 65 years and older) and individuals with underlying conditions such as cancer, diabetes, or liver diseases^9^. The Centers for Disease Control and Prevention (CDC) state that individuals of any age with underlying conditions are at higher risk of developing severe symptoms from COVID-19^10^. Moreover, UNICEF warns that children with underlying disabilities may be at greater risk of developing complications^11^. More data, including the presence of brain-based disabilities and other at-risk conditions in children, are required to have a better understanding of the clinical impacts of COVID-19 on these potentially more vulnerable populations.

### Objectives

This rapid review was commissioned by the Strategy for Patient-Oriented Research (SPOR) funded CHILD-BRIGHT Network, an innovative pan-Canadian research network that aims to improve life outcomes for children with brain-based developmental disabilities and their families (https://www.child-bright.ca). Concerned with the potential impact of the novel coronavirus on children with brain-based developmental disabilities, they requested support from the SPOR Evidence Alliance to conduct a rapid review on the topic. Thus, this review aimed to answer the following questions:

1. Are children with brain-based developmental disabilities more likely to develop COVID-19?
2. Are children with brain-based developmental disabilities more likely to develop complications due to COVID-19?
3. Are children with brain-based developmental disabilities more likely to have a poorer prognosis once they develop COVID-19?

We engaged with a panel of knowledge users (patients, caregivers, clinicians, decision makers) and researchers from the CHILD-BRIGHT Network throughout the review process, from question development, literature search, interpretation, writing of results, to dissemination of findings.

## Methods

We conducted a rapid review based on the proposed methodology guide of the Cochrane Rapid Reviews Methods Group^12^. We report our results based on the Preferred Reporting Items for Systematic Reviews and Meta-Analyses (PRISMA) Statement^13^.

### Literature Search

An experienced medical information specialist developed the search strategies through an iterative process in consultation with the review team and the panel of knowledge users. The MEDLINE strategy was peer reviewed by another senior information specialist prior to execution using the PRESS Checklist^14^. Using the OVID platform, we searched Ovid MEDLINE®, including Epub Ahead of Print and In-Process & Other Non-Indexed Citations, Embase Classic+Embase, PsycINFO, Cochrane Database of Systematic Reviews and the Cochrane Central Register of Controlled Trials. We also searched CINAHL (Ebsco) and Web of Science. Searches were initially performed on April 18th, 2020^15^, and updated on October 31st, 2020.

We used a combination of controlled vocabulary (e.g., “Coronavirus Infections”, “Child”, “Pediatrics”) and keywords (e.g., “coronavirus”, “COVID-19”, “infant”) for the strategies and adjusted vocabulary and syntax across databases. There were no language restrictions on any of the searches but when possible, animal-only records were removed from the results. We limited results to the publication years 2019 to the present. Due to the increase in volume of Covid-19 related literature, the updated search incorporated vocabulary pertaining to specific brain, developmental and substance abuse disorders (this terminology was included in the original peer review assessment). Specific details regarding the strategies appear in Appendix 1.

We also performed grey literature searches, which consisted of preprint articles from SSRN and MedRxiv (last consulted November 13th, 2020), ongoing trials from the WHO International Clinical Trials Registry Platform (last consulted November 13th, 2020), ongoing reviews from PROSPERO (last consulted November 13th, 2020), and Government or Health organizations’ websites and reports (consulted between November 5th to November 13th, 2020) (Appendix 2).

### Eligibility Criteria

We followed the PECO Framework in establishing eligibility criteria^16, 17^ (Table 1). We considered any study with primary data that included children aged between 0 and 18 with a brain-based developmental disability or at risk of developing such disability with confirmed or suspected COVID-19 (see Appendix 3 for a list of eligible conditions).

**Table 1.** PECO Inclusion Criteria.

|  |  |
| --- | --- |
| **Population (P)** | Children (18 years and under) with brain-based developmental disabilities (e.g., cerebral palsy, autism, developmental delay, ADHD, severe impairments) or at-risk of developing brain-based disabilities (e.g., premature, congenital heart disease/defect). |
| **Exposure (E)** | COVID-19 |
| **Comparator (C)** | Any or none |
| **Outcomes (O)** | Any outcome |

### Study Selection and Extraction

Four reviewers individually performed screening for titles, abstracts and then full text using pilot-tested standardized forms. We developed a standardized extraction form that included study characteristics (e.g., authors, country, study design), patients’ characteristics (e.g., condition, sex, age), and outcomes reported. Single reviewers extracted the data.

### Quality Assessment

We used the Joanna Briggs Institute (JBI) Critical Appraisal Checklists to assess the methodological quality of included studies^18^. Due to the diversity of designs included in this review, we used three different evaluation grids: one for cohort studies, one for case reports, and one for case series. Each grid evaluates eight to 11 key biases relevant to each study design (see Appendix 4 for the list of quality indicators). Type of answers is the same for each grid: yes, no, unclear, not applicable. Two reviewers individually assessed the quality.

### Synthesis

We report data using a narrative approach which includes tables of study characteristics, and detailed reporting of patients’ characteristics, treatments, and outcomes. Our data synthesis focused on providing a descriptive summary to inform our knowledge users.

## Results

### Literature Search

Our bibliographic database searches led to the identification of 1446 deduped publications, from which 333 were assessed for eligibility at full text. In total, 29 articles were included in this review (Figure 1)^19-47^.

**Figure 1.**
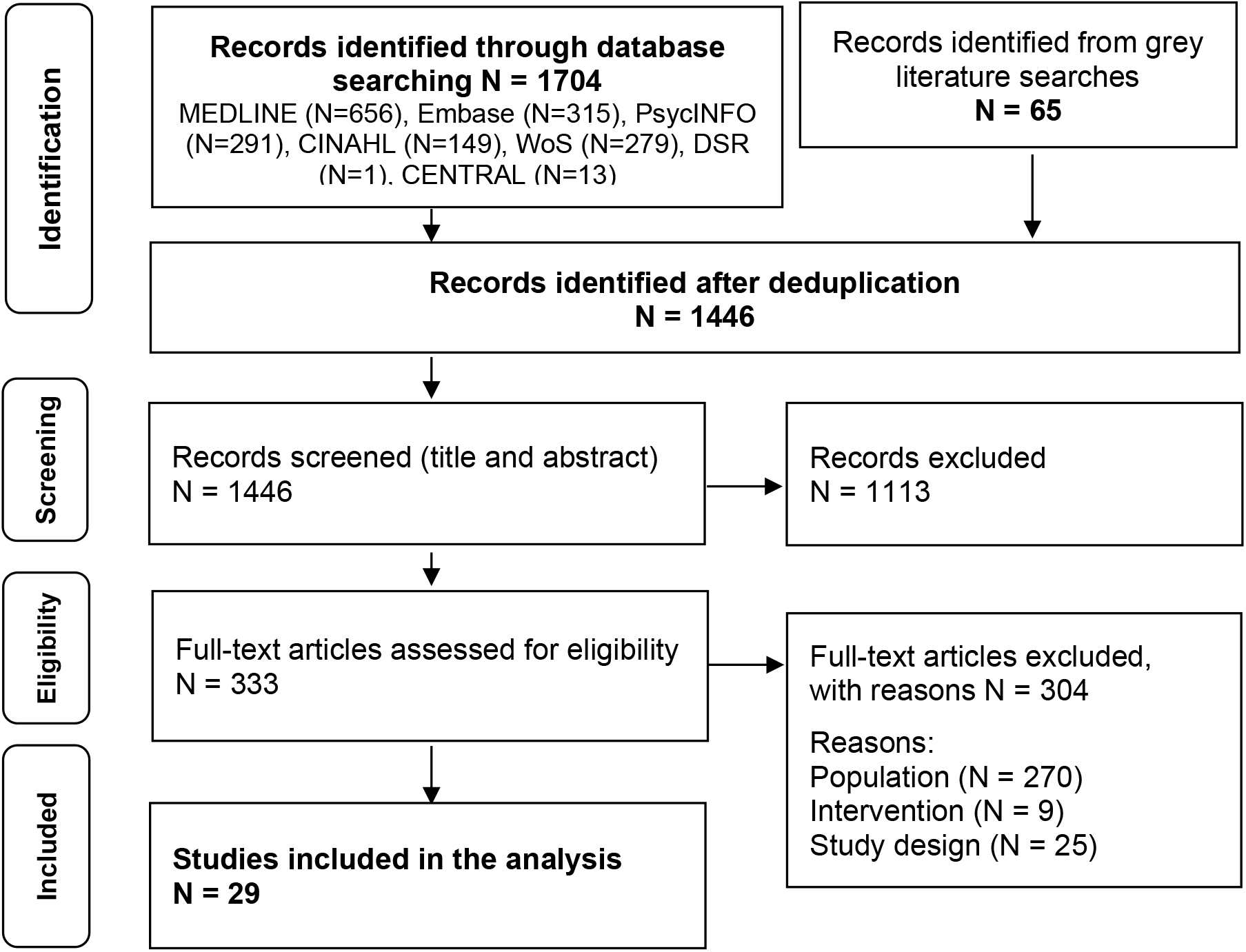
Study Flow Diagram.

### Characteristics of Included Studies

Nine studies^21, 24, 27, 29, 31, 33, 36, 39, 42^ included children with brain-based disabilities, and the remaining 20 included children at risk of developing a brain-based disability (premature infants and infants with congenital heart diseases in most studies) (Table 2). Most studies were published in the United States and the majority of studies were case reports or case series (23/29, 79%).

**Table 2.**
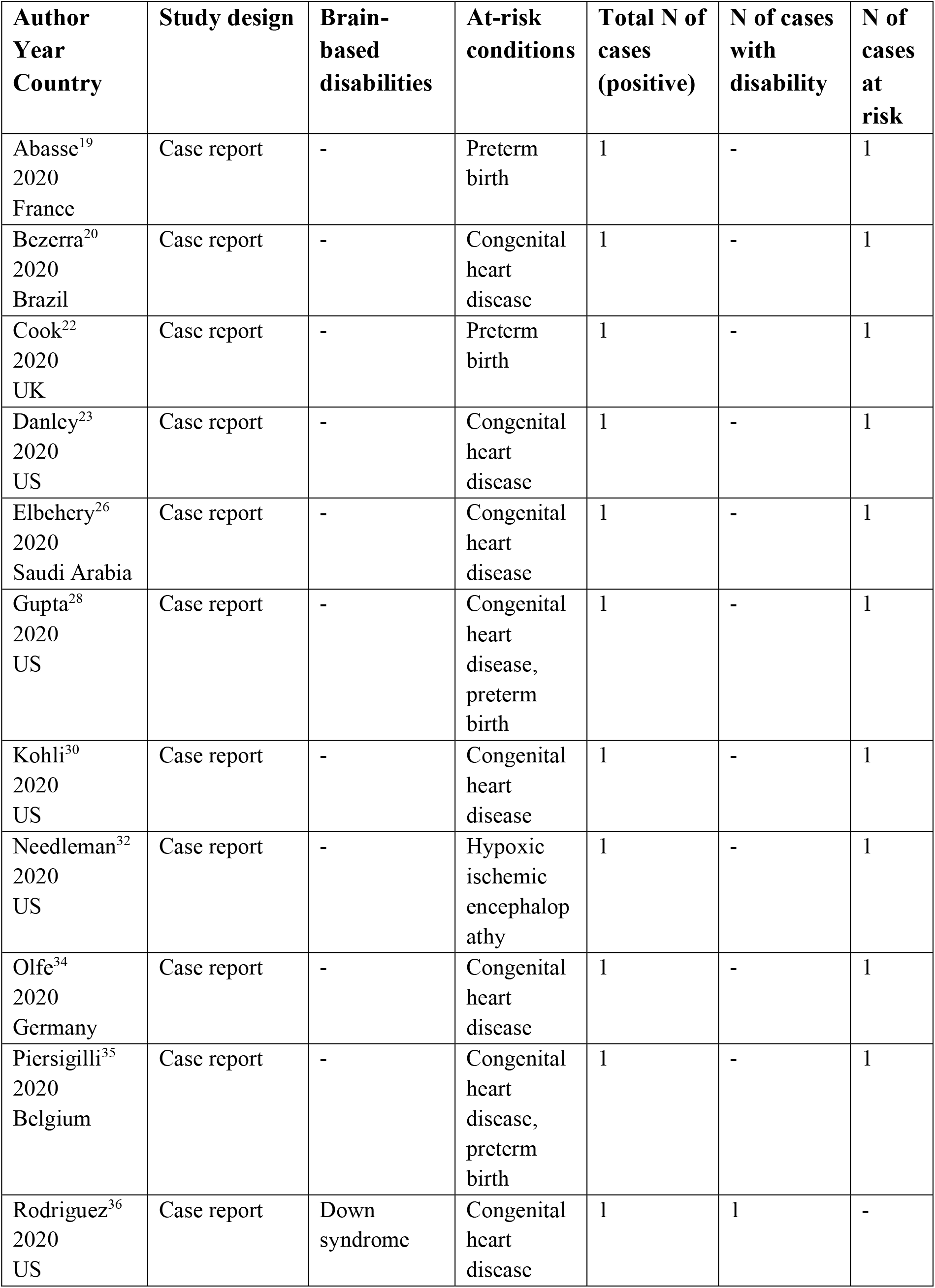
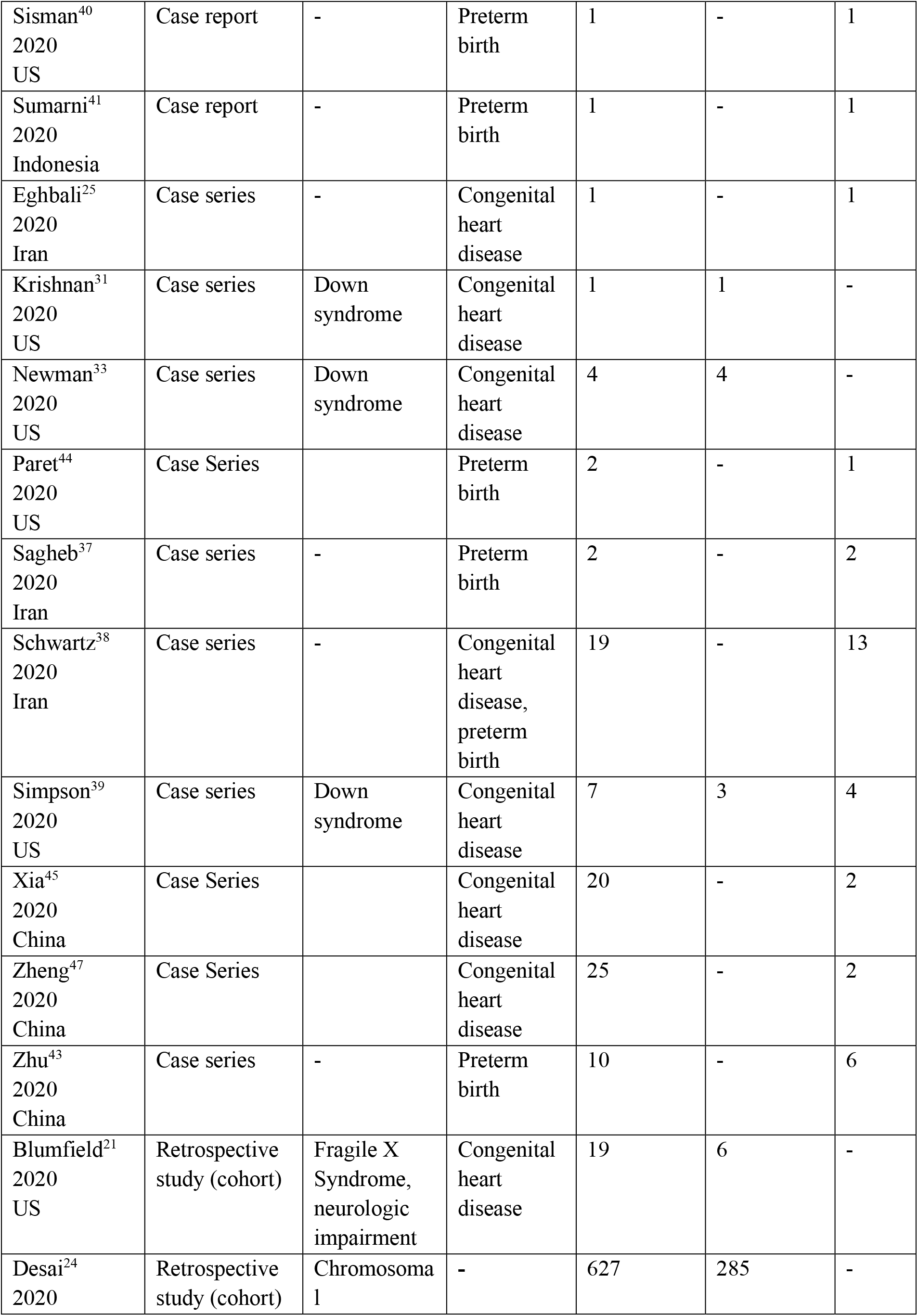
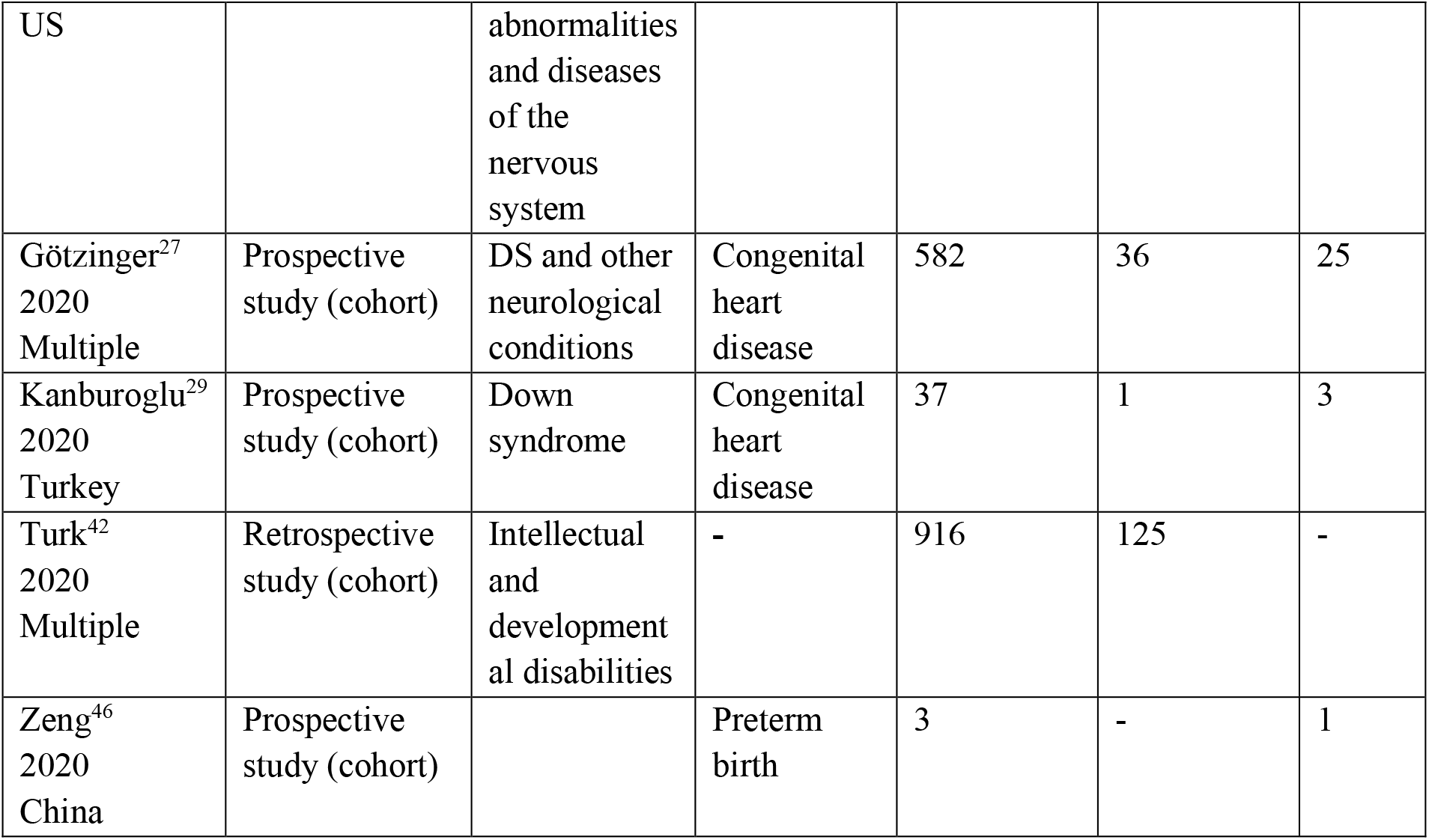
Description of included studies.

### Quality of Included Studies

Of the 13 case reports, 95% of responses to quality assessment questions were positive (Appendix 4). All studies met the first three quality criteria, which focused on the description of patients’ demographic characteristics, history and clinical condition. Three out of 13 case reports did not fully report diagnostic tests or assessment methods.

Of the 10 case series reports, the average of positive responses to quality assessment questions was 79%. For most of the case series, information regarding the inclusion of participants was not available, particularly in terms of the consecutive and complete nature of the inclusion of participants.

Of the six cohort studies analyzed, 90% of responses to quality assessment questions were positive. We modified the interpretation of the first criteria on exposure since all cohort studies included in this review only had a single group of COVID positive patients. Also, the criteria on confounding factors were not applicable since COVID-19 is an emerging disease, which does not allow us to fully identify the confounding factors that are associated with it. In all cohort studies, no case was lost during follow-up.

### Patient Characteristics

The included studies reported data on 2288 COVID-19 positive children, including 462 with a brain-based disability, and 72 at risk of developing such disability (Table 3 and 4). The mean age of children with brain-based disabilities or at risk for disability was four years old and 48% were female. Most conditions were classified in broad categories, such as neurological disorders or chromosomal abnormalities. Forty children had a congenital heart disease, and 29 were premature infants. More than half of the children with Down syndrome also had a congenital heart disease.

**Table 3.** Number of cases in children with brain-based disabilities.

| **Number of cases** | **Brain-based disabilities** |
| --- | --- |
| 26 | Neurological disorders |
| 18* | Down syndrome |
| 5 | Neurologic impairments |
| 1** | Fragile X syndrome |
| 125 | Intellectual and developmental disabilities |
| 138 | Chromosomal abnormalities |
| 149 | Diseases of the nervous system |
| **462** | **Total number of cases** |

\*Ten children with Down syndrome also had a congenital heart disease.
\*\*One child with Fragile X syndrome also had a congenital heart disease.

**Table 4.**
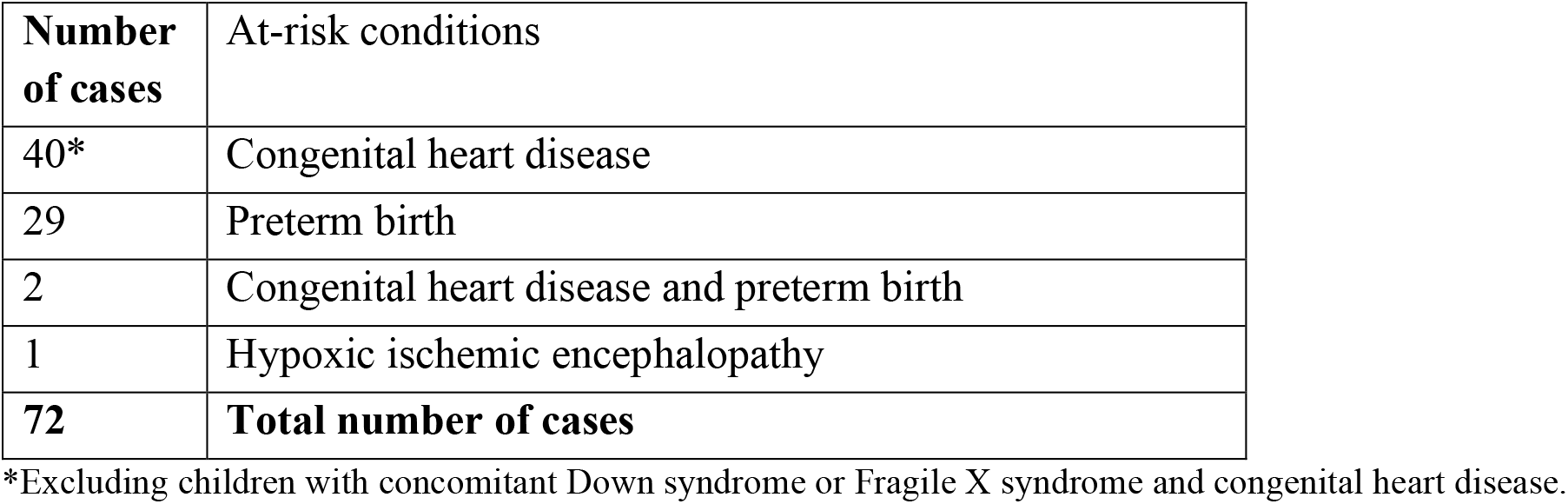
Number of cases in children at risk of developing brain-based disabilities.

For children with or at risk for brain-based disabilities, whose COVID-19 symptoms were reported (n=50), the most frequent were breathing issues (56%), fever (48%), hypoxemia (20%), decreased oral intake (20 %) and cough (18%). For children with Down syndrome for whom symptoms were reported (n=9), 67% had fever, 56% had cough and 56% had breathing issues.

Antibiotics were the most frequently reported medication (27 studies, 93%), followed by antiviral medications (15 studies, 52%), and anti-inflammatory drugs (11 studies, 38%). Hydroxychloroquine was reported in eight studies (28%). Oxygen delivered though a nasal cannula was the most frequently reported treatment (22 studies, 76%), followed by mechanical ventilation (15 studies, 41%), and non-invasive pressure ventilation (six studies, 21%).

### Case Reports and Case Series

The 23 case reports and case series included a total of 52 children with brain-based disability or at risk of developing such a disability, amongst which there were nine children with Down syndrome, 26 with congenital heart disease, and 27 preterm infants (non-mutually exclusive).

Authors from the case reports and case series reported several children that required admittance to the Intensive Care Unit (ICU). In children with Down syndrome, 67% (six out of nine) of them were admitted to the ICU. Also, children with comorbidities were often admitted to ICUs. In children with congenital heart disease or prematurity, 58% (15/26) and 67% (18/27) of them respectively were admitted to ICU.

The most common type of complication observed in children included in the case reports and case series was acute respiratory distress syndrome (Table 5, 6, 7). More specifically, three children with Down syndrome, nine children with congenital heart disease and nine premature infants suffered from acute respiratory distress. The second most common complication in these children was pneumonia, affecting two children with Down syndrome, five children with congenital heart disease and four premature infants. Other complications included heart failure (two cases with Down syndrome, five cases with congenital heart disease), bacterial or viral co-infection (two cases with Down syndrome, three cases with congenital heart disease), pneumothorax (one case with congenital heart disease, two premature infants) and acute renal injury (one case with Down syndrome, three cases with congenital heart disease). Among all cases reported, seven children died: two with Down syndrome, one with congenital heart disease, and four premature infants. Several children did not experience any complications (five out of nine children with Down syndrome, nine of 26 children with congenital heart disease, and 13 of 27 preterm infants).

**Table 5.**
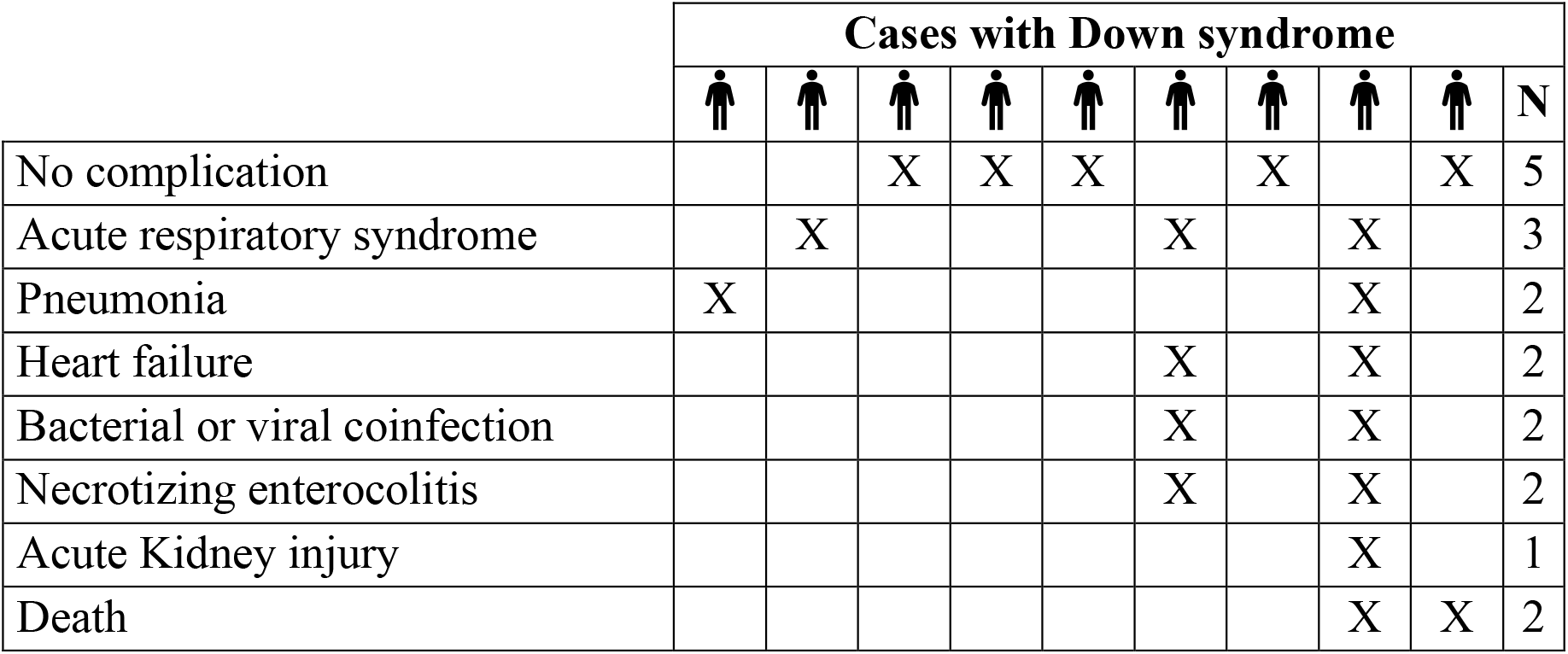
Complications for each child with Down syndrome.

|  | Cases with Down syndrome |  |  |  |  |  |  |  |  |  |
| --- | --- | --- | --- | --- | --- | --- | --- | --- | --- | --- |
| No complication |  |  | X | X | X |  | X |  | X | 5 |
| Acute respiratory syndrome |  | X |  |  |  | X |  | X |  | 3 |
| Pneumonia | X |  |  |  |  |  |  | X |  | 2 |
| Heart failure |  |  |  |  |  | X |  | X |  | 2 |
| Bacterial or viral coinfection |  |  |  |  |  | X |  | X |  | 2 |
| Necrotizing enterocolitis |  |  |  |  |  | X |  | X |  | 2 |
| Acute Kidney injury |  |  |  |  |  |  |  | X |  | 1 |
| Death |  |  |  |  |  |  |  | X | X | 2 |

**Table 6.**
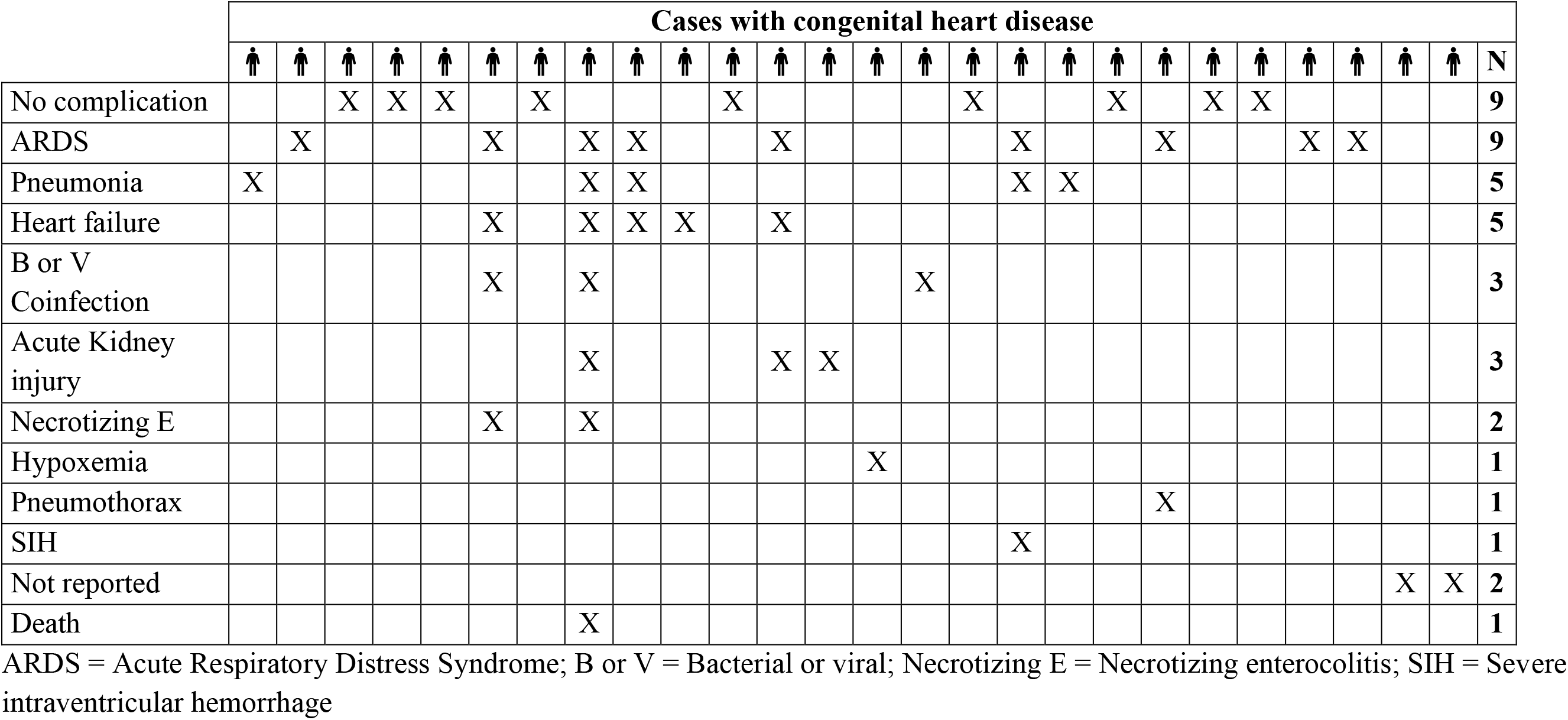
Complications for each child with congenital heart disease.

**Table 7.**
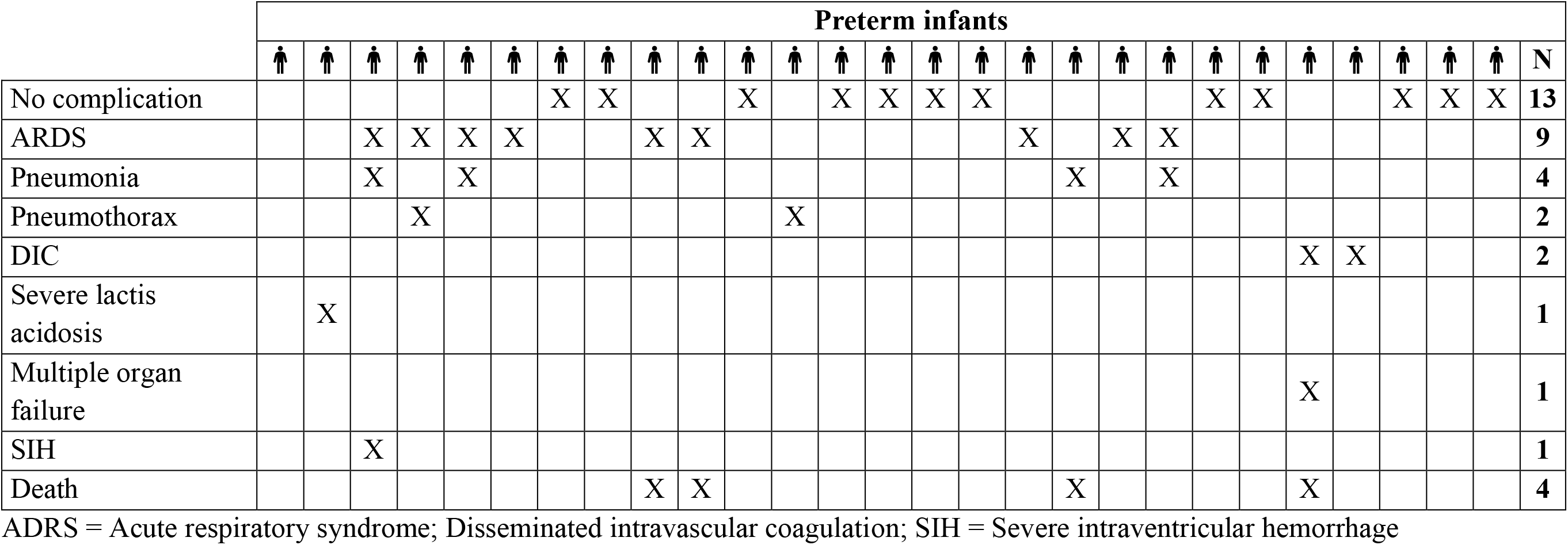
Complications for each preterm infant.

### Cohort Studies

A total of six cohort studies reported on 451 children with brain-based developmental disabilities and 29 at-risk children.

Zeng and colleagues ^46^ examined the occurrence of COVID-19 transmission from mothers to newborns in Wuhan, China. Of the 33 infants included in the study, only three were infected, from which one was born prematurely. The child had severe complications following birth (respiratory distress and pneumonia) which required resuscitation. The infant was discharged after 11 days in intensive care.

Blumfield et al.^21^ retrospectively reviewed the medical records of 19 children and adolescents with COVID-19 admitted to the hospital in New York City. Twelve patients had comorbidities, including five who had severe neurologic impairments and one with Fragile X syndrome and congenital heart disease. Among these 19 patients, 14 were admitted to intensive care and two, with unspecified comorbidities, died.

In the study by Kanburoglu et al^29^, 37 newborns from Turkey who contracted COVID-19 in the community and were admitted to intensive care were enrolled in the study. Of these infants, one had Down syndrome and congenital heart disease, and three were premature infants. The child with Down syndrome died after 21 days in intensive care from acute respiratory distress and bacterial co-infection. The mean hospitalization length for all cases was 11 days, ranging from one to 35 days.

Desai and colleagues^24^ examined electronic medical records of 627 children with COVID-19 in the United States. Of the 293 children hospitalized, 28 (10%) were considered severe. Of these severe cases, at least 20 had a disability. Hospitalized children were younger than those not hospitalized (5.6 years versus (vs) 8.2 years, p<0.001). Among the severe cases, there were more children with underlying conditions, including chromosomal abnormalities (71% vs 22%, p <0.001), as well as children with a nervous system disease (71% vs 23%, p <0.001). This was also the case in children with heart problems (50% vs 11%, p <0.001) and other comorbidities. One patient died, but there were no reported details related to the child’ pre-existing conditions.

Götzinger et al^27^ presented the analysis of 582 children with COVID-19 in Europe. A total of 363 (62%) children were hospitalized and 48 (8%) were admitted to intensive care. Among children with comorbidities (n=145, 25% of all cases), 26 had neurological disorders, 25 had congenital heart diseases, and 10 had chromosomal abnormalities. Univariate analyzes demonstrated that the odds to be admitted to intensive care were higher in children with neurologic disorders and congenital heart diseases (respectively, odds ratio [OR]=2.8, 95% CI 1.0-7.9, p=0.037, and OR=2.9, 95% CI 1.0-8.4, p=0.029). Results were not statistically significant in children with chromosomal abnormalities (OR=2.8, 95% CI 0.5-13.8, p=0.19). For multivariate analyses, OR for all pre-existing conditions combined was 3.27 (95% CI 1.67-6.42, p=0.0015). Authors also pointed out that children less than one month old were more at risk to be admitted to intensive care (OR=5.06, 95% CI 1.72-14.87, p=0.0035). During the study, four children died, including two with comorbidities, without further details by the authors. The reported case-fatality rate was 0.69% (95% CI 0.20-1.82).

Turk and colleagues^42^ analyzed thousands of electronic medical records to compare differences in deaths and comorbidities between patients of all ages with and without intellectual and developmental disabilities. Their resulting database included more than 30,000 patients with COVID-19, including 916 children aged 17 and under, of which 125 had a disability. For all cases with disabilities, 26.4% were children (125/474). In comparison, for all cases without disabilities, 2.7% were children (791/29808). The case-mortality rate was similar for patients with or without disability for all ages (around 5%). However, differences were identified between age groups. In children aged 17 years old or less, the case-fatality rate was higher in children with disability than without (1.6%, 95% CI 0.4-5.6, vs. 0.1%, 95% CI 0.0-0.7). This difference persisted for adults between the ages of 18 and 75 (4.5%, 95% CI 2.7-7.4, vs. 2.7%, 95% CI 2.5-3.0) but did not persist for people aged 75 or more (21.1%, 95% CI 11.1-36.3, vs. 20.7%, 95% CI 19.5-21.9).

## Discussion

Overall, the included studies showed that there is a greater risk of developing severe COVID-19 disease in children with brain-based disabilities, with pre-existing comorbid conditions, and in younger children.

Although mortality is very low among children, the case-fatality rate appears to be higher in children with disabilities, compared to children without disabilities^42^. Since comorbidities are more frequent in individuals with brain-based disabilities, including children, this may partly explain their higher risk of developing severe COVID-19 disease. As highlighted by Desai^24^ and Götzinger^27^, younger age may also play a role in illness severity.

Many studies (e.g. Götzinger^27^, Blumfied^21^, and Kanburoglu^29^) included children that were hospitalized due to COVID-19, often in secondary and tertiary care centers. Results of this review may therefore seem alarming, but this does not capture information about children with disabilities who have contracted COVID-19 but have not been hospitalized. Furthermore, illness severity in children seems generally low; in Canada, as of November 25, 2020, only 1.4% of the hospitalized cases were children, with two reported deaths^2^.

Additionally, case-fatality rates reported in the included studies were calculated on a relatively low number of children with disabilities, which may question their reliability. They were also limited to the cases analysed by the studies, which may affect generalizability. Case-fatality rates vary significantly from one country to another, and may be influenced by several factors, such as country age distribution, testing strategies, and definition of COVID-19-related deaths. Also, case-fatality rates should be interpreted with caution, because asymptomatic and undiagnosed cases may fall under the radar.

Our rapid review had certain limitations. Due to our study design, screening and data extraction were performed by single reviewers. However, we did perform pilot-testing for each review form to optimize consistency between reviewers. Despite our best efforts to identify all relevant studies, it is possible that some were missed. To mitigate this, we searched multiple grey literature sources, including preprint databases, review and trial registries, and several websites.

## Data Availability

Data is available upon request.

## Acknowledgements

The authors would like to thank the panel of knowledge users for their support throughout this review. This project was funded by the CHILD-BRIGHT Network and the SPOR Evidence Alliance. Both Networks are supported by the Canadian Institutes of Health Research under Canada’s Strategy for Patient-Oriented Research (SPOR) Initiative.

## Conflict of Interest Disclosures

The authors declare that they have no conflict of interests.

## Funding

This review was funded by the CHILD-BRIGHT Network (https://www.child-bright.ca) and the SPOR Evidence Alliance. Both Networks are supported by the Canadian Institutes of Health Research (CIHR) under Canada’s Strategy for Patient-Oriented Research (SPOR) Initiative.

## Notes

### Competing Interest Statement

The authors have declared no competing interest.

### Author Declarations

This is a rapid review. No IRB needed.

