## Supplemental Data 1 for "COVID-19 in Children with Brain-Based Developmental Disabilities: A Rapid Review Update"

**Appendix 1. Search Strategies**

2020 Apr 18

Ovid Multifile

Database: Ovid MEDLINE: Epub Ahead of Print, In-Process & Other Non-Indexed Citations, Ovid MEDLINE® Daily and Ovid MEDLINE® <1946-Present>, Embase Classic+Embase <1947 to 2020 April 16>, APA PsycInfo <1806 to April Week 2 2020>

Search Strategy:

--------------------------------------------------------------------------------

1	Coronavirus/ (9006)

2	Coronavirus Infections/ (6788)

3	(COVID-19 or COVID19).mp. (7538)

4	((coronavirus* or corona virus*) and (hubei or wuhan or beijing or shanghai)).mp. (1621)

5	Wuhan virus*.mp. (12)

6	2019-nCoV.mp. (1049)

7	(nCoV or n-CoV).mp. (1098)

8	HCoV-19.mp. (8)

9	(SARS-CoV-2 or SARS-CoV2 or SARSCoV-2 or SARSCoV2).mp. (2406)

10	(novel coronavirus* or novel corona virus*).mp. (3086)

11	((coronavirus* or corona virus*) adj2 "2019").mp. (2881)

12	((coronavirus* or corona virus*) adj2 "19").mp. (533)

13	(coronavirus 2 or corona virus 2).mp. (1942)

14	(coronavirus* or corona virus*).ti. (16258)

15	or/1-14 [COVID 19] (29699)

16	exp Infant/ (2278037)

17	exp Child/ (4879551)

18	Adolescent/ (3645659)

19	(baby or babies or infant? or infanc* or neonat* or newborn* or preschool* or pre-school* or toddler?).tw,kf. (1924540)

20	(adolescen* or child* or pre-adolescen* or preteen* or pre-teen* or school-age* or teen or teens or teenager? or youth*).tw,kf. (4626733)

21	exp Pediatrics/ (203872)

22	p?ediatric*.tw,kf. (943730)

23	or/16-22 [INFANTS/CHILDREN/ADOLESCENTS] (9782583)

24	15 and 23 [COVID-19 - INFANTS/CHILDREN/ADOLESCENTS] (2990)

25	exp Animals/ not Humans/ (18169986)

26	24 not 25 [ANIMAL-ONLY REMOVED] (2013)

27	limit 26 to yr="2019-current" (658)

28	27 use ppez [MEDLINE RECORDS] (386)

29	Coronavirinae/ (3474)

30	Coronavirus infection/ (7716)

31	(COVID-19 or COVID19).mp. (7538)

32	((coronavirus* or corona virus*) and (hubei or wuhan or beijing or shanghai)).mp. (1621)

33	Wuhan virus*.mp. (12)

34	2019-nCoV.mp. (1049)

35	(nCoV or n-CoV).mp. (1098)

36	HCoV-19.mp. (8)

37	(SARS-CoV-2 or SARS-CoV2 or SARSCoV-2 or SARSCoV2).mp. (2406)

38	(novel coronavirus* or novel corona virus*).mp. (3086)

39	((coronavirus* or corona virus*) adj2 "2019").mp. (2881)

40	((coronavirus* or corona virus*) adj2 "19").mp. (533)

41	(coronavirus 2 or corona virus 2).mp. (1942)

42	(coronavirus* or corona virus*).ti. (16258)

43	or/29-42 [COVID 19] (27586)

44	exp child/ (4879551)

45	exp adolescent/ (3645863)

46	juvenile/ (49134)

47	(baby or babies or infant? or infanc* or neonat* or newborn* or preschool* or pre-school* or toddler?).tw,kw. (1906006)

48	(adolescen* or child* or pre-adolescen* or preteen* or pre-teen* or school-age* or teen or teens or teenager? or youth*).tw,kw. (4632750)

49	exp pediatrics/ (203872)

50	p?ediatric*.tw,kw. (969422)

51	or/44-50 [INFANTS/CHILDREN/ADOLESCENTS] (9624446)

52	43 and 51 [COVID-19 - INFANTS/CHILDREN/ADOLESCENTS] (2521)

53	exp animal/ or exp animal experimentation/ or exp animal model/ or exp animal experiment/ or nonhuman/ or exp vertebrate/ (52820606)

54	exp human/ or exp human experimentation/ or exp human experiment/ (40547950)

55	53 not 54 (12274383)

56	52 not 55 [ANIMAL-ONLY REMOVED] (1987)

57	limit 56 to yr="2019-current" (734)

58	57 use emczd [EMBASE RECORDS] (351)

59	(COVID-19 or COVID19).mp. (7538)

60	((coronavirus* or corona virus*) and (hubei or wuhan or beijing or shanghai)).mp. (1621)

61	Wuhan virus*.mp. (12)

62	2019-nCoV.mp. (1049)

63	(nCoV or n-CoV).mp. (1098)

64	HCoV-19.mp. (8)

65	(SARS-CoV-2 or SARS-CoV2 or SARSCoV-2 or SARSCoV2).mp. (2406)

66	(novel coronavirus* or novel corona virus*).mp. (3086)

67	((coronavirus* or corona virus*) adj2 "2019").mp. (2881)

68	((coronavirus* or corona virus*) adj2 "19").mp. (533)

69	(coronavirus 2 or corona virus 2).mp. (1942)

70	(coronavirus* or corona virus*).ti. (16258)

71	or/59-70 [COVID 19] (23023)

72	(baby or babies or infant? or infanc* or neonat* or newborn* or preschool* or pre-school* or toddler?).tw,id. (1874620)

73	(adolescen* or child* or pre-adolescen* or preteen* or pre-teen* or school-age* or teen or teens or teenager? or youth*).tw,id. (4573083)

74	exp Pediatrics/ (203872)

75	p?ediatric*.tw,id. (928599)

76	limit 71 to (100 childhood <birth to age 12 yrs> or 120 neonatal <birth to age 1 mo> or 140 infancy <2 to 23 mo> or 160 preschool age <age 2 to 5 yrs> or 180 school age <age 6 to 12 yrs> or 200 adolescence <age 13 to 17 yrs>) [Limit not valid in Ovid MEDLINE(R),Ovid MEDLINE(R) Daily Update,Ovid MEDLINE(R) In-Process,Ovid MEDLINE(R) Publisher,Embase; records were retained] (22978)

77	72 or 73 or 74 or 75 (6217215)

78	71 and 77 (1436)

79	76 or 78 [COVID-19 - INFANT/CHILD/ADOLESCENT POPULATION] (22980)

80	limit 79 to yr="2019-current" (10272)

81	80 use ppez,emczd (10272)

82	80 not 81 [PSYCINFO RECORDS] (0)

83	28 or 58 or 82 [ALL DATABASES] (737)

84	remove duplicates from 83 (485)

85	84 use ppez [MEDLINE UNIQUE RECORDS] (367)

86	84 use emczd [EMBASE UNIQUE RECORDS] (118)

87	84 not (85 or 86) [PSYCINFO UNIQUE RECORDS] (0)

***************************

CINAHL

| # | Query | Limiters/Expanders | Last Run Via | Results |
| --- | --- | --- | --- | --- |
| S17 | S9 AND S15 | Limiters - Published Date: 20190101-20201231  Search modes - Boolean/Phrase | Interface - EBSCOhost Research Databases  Search Screen - Advanced Search  Database - CINAHL Plus with Full Text | 85 |
| S16 | S9 AND S15 | Search modes - Boolean/Phrase | Interface - EBSCOhost Research Databases  Search Screen - Advanced Search  Database - CINAHL Plus with Full Text | 392 |
| S15 | S10 OR S11 OR S12 OR S13 OR S14 | Search modes - Boolean/Phrase | Interface - EBSCOhost Research Databases  Search Screen - Advanced Search  Database - CINAHL Plus with Full Text | 1,290,363 |
| S14 | TI p#ediatric* OR AB p#ediatric* | Search modes - Boolean/Phrase | Interface - EBSCOhost Research Databases  Search Screen - Advanced Search  Database - CINAHL Plus with Full Text | 141,058 |
| S13 | (MH "Pediatrics+") | Search modes - Boolean/Phrase | Interface - EBSCOhost Research Databases  Search Screen - Advanced Search  Database - CINAHL Plus with Full Text | 22,532 |
| S12 | TI ( adolescen* or child* or pre-adolescen* or preteen* or pre-teen* or school-age* or teen or teens or teenager# or youth* ) OR AB ( adolescen* or child* or pre-adolescen* or preteen* or pre-teen* or school-age* or teen or teens or teenager# or youth* ) | Search modes - Boolean/Phrase | Interface - EBSCOhost Research Databases  Search Screen - Advanced Search  Database - CINAHL Plus with Full Text | 638,958 |
| S11 | TI ( baby or babies or infant# or infanc* or neonat* or newborn* or preschool* or pre-school* or toddler# ) OR AB ( baby or babies or infant# or infanc* or neonat* or newborn* or preschool* or pre-school* or toddler# ) | Search modes - Boolean/Phrase | Interface - EBSCOhost Research Databases  Search Screen - Advanced Search  Database - CINAHL Plus with Full Text | 211,822 |
| S10 | (MH "Child+") OR (MH "Minors (Legal)") OR (MH "Adolescence") | Search modes - Boolean/Phrase | Interface - EBSCOhost Research Databases  Search Screen - Advanced Search  Database - CINAHL Plus with Full Text | 1,030,685 |
| S9 | S1 OR S2 OR S3 OR S4 OR S5 OR S6 OR S7 OR S8 | Search modes - Boolean/Phrase | Interface - EBSCOhost Research Databases  Search Screen - Advanced Search  Database - CINAHL Plus with Full Text | 3,134 |
| S8 | TI coronavirus* or (corona N0 virus*) | Search modes - Boolean/Phrase | Interface - EBSCOhost Research Databases  Search Screen - Advanced Search  Database - CINAHL Plus with Full Text | 856 |
| S7 | TX ( (novel N0 coronavirus*) or ("novel corona" N0 virus*) ) OR TX ( (coronavirus* or (corona" N0 virus*)) N2 "2019" ) OR TX ( (coronavirus* or (corona" N0 virus*)) N2 "19" ) OR TX ( "coronavirus 2" or "corona virus 2" ) | Search modes - Boolean/Phrase | Interface - EBSCOhost Research Databases  Search Screen - Advanced Search  Database - CINAHL Plus with Full Text | 2,761 |
| S6 | TX "2019-nCoV" OR TX ( nCoV or "n-CoV" ) OR TX "HCoV-19" OR TX ( "SARS-CoV-2" or "SARS-CoV2" or "SARSCoV-2" or SARSCoV2 ) | Search modes - Boolean/Phrase | Interface - EBSCOhost Research Databases  Search Screen - Advanced Search  Database - CINAHL Plus with Full Text | 218 |
| S5 | TX Wuhan N0 virus* | Search modes - Boolean/Phrase | Interface - EBSCOhost Research Databases  Search Screen - Advanced Search  Database - CINAHL Plus with Full Text | 3 |
| S4 | TX ( coronavirus* and (hubei or wuhan or beijing or shanghai) ) OR TX ( (corona N0 virus*) and (hubei or wuhan or beijing or shanghai) ) | Search modes - Boolean/Phrase | Interface - EBSCOhost Research Databases  Search Screen - Advanced Search  Database - CINAHL Plus with Full Text | 307 |
| S3 | TX "COVID-19" or COVID19 | Search modes - Boolean/Phrase | Interface - EBSCOhost Research Databases  Search Screen - Advanced Search  Database - CINAHL Plus with Full Text | 819 |
| S2 | (MH "Coronavirus Infections") | Search modes - Boolean/Phrase | Interface - EBSCOhost Research Databases  Search Screen - Advanced Search  Database - CINAHL Plus with Full Text | 981 |
| S1 | (MH "Coronavirus") | Search modes - Boolean/Phrase | Interface - EBSCOhost Research Databases  Search Screen - Advanced Search  Database - CINAHL Plus with Full Text | 302 |

Cochrane

Database: EBM Reviews - Cochrane Central Register of Controlled Trials <March 2020>, EBM Reviews - Cochrane Database of Systematic Reviews <2005 to April 17, 2020>

Search Strategy:

--------------------------------------------------------------------------------

1	Coronavirus/ (2)

2	Coronavirus Infections/ (47)

3	(COVID-19 or COVID19).mp. (38)

4	((coronavirus* or corona virus*) and (hubei or wuhan or beijing or shanghai)).mp. (23)

5	Wuhan virus*.mp. (0)

6	2019-nCoV.mp. (25)

7	(nCoV or n-CoV).mp. (26)

8	HCoV-19.mp. (0)

9	(SARS-CoV-2 or SARS-CoV2 or SARSCoV-2 or SARSCoV2).mp. (10)

10	(novel coronavirus* or novel corona virus*).mp. (30)

11	((coronavirus* or corona virus*) adj2 "2019").mp. (25)

12	((coronavirus* or corona virus*) adj2 "19").mp. (6)

13	(coronavirus 2 or corona virus 2).mp. (2)

14	(coronavirus* or corona virus*).ti. (43)

15	or/1-14 [COVID 19] (105)

16	exp Infant/ (30812)

17	exp Child/ (54451)

18	Adolescent/ (102852)

19	(baby or babies or infant? or infanc* or neonat* or newborn* or preschool* or pre-school* or toddler?).ti,ab,kw. (69335)

20	(adolescen* or child* or pre-adolescen* or preteen* or pre-teen* or school-age* or teen or teens or teenager? or youth*).ti,ab,kw. (167037)

21	exp Pediatrics/ (653)

22	p?ediatric*.ti,ab,kw. (34019)

23	or/16-22 (292919)

24	15 and 23 [COVID - PAEDIATRIC POPULATION] (13)

25	limit 24 to yr="2019-current" (4)

26	25 use coch [DSR RECORDS] (0)

27	25 use cctr [CENTRAL RECORDS] (4)

***************************

Web of Science

| # 7 | [78](https://apps-webofknowledge-com.myaccess.library.utoronto.ca/summary.do?product=WOS&doc=1&qid=32&SID=5ElQLmIsRtNxI8dydUD&search_mode=CombineSearches&update_back2search_link_param=yes) | #5 AND #4  Refined by: PUBLICATION YEARS: ( 2020 OR 2019 )  Indexes=SCI-EXPANDED, SSCI, A&HCI, CPCI-S, CPCI-SSH, BKCI-S, BKCI-SSH, ESCI Timespan=All years |
| --- | --- | --- |
| # 6 | [552](https://apps-webofknowledge-com.myaccess.library.utoronto.ca/summary.do?product=WOS&doc=1&qid=31&SID=5ElQLmIsRtNxI8dydUD&search_mode=CombineSearches&update_back2search_link_param=yes) | #5 AND #4  Indexes=SCI-EXPANDED, SSCI, A&HCI, CPCI-S, CPCI-SSH, BKCI-S, BKCI-SSH, ESCI Timespan=All years |
| # 5 | [2,820,912](https://apps-webofknowledge-com.myaccess.library.utoronto.ca/summary.do?product=WOS&doc=1&qid=21&SID=5ElQLmIsRtNxI8dydUD&search_mode=GeneralSearch&update_back2search_link_param=yes) | TOPIC: (baby or babies or infant or infants or infanc* or neonat* or newborn* or preschool* or pre-school* or toddler*) OR TOPIC: (adolescen* or child* or pre-adolescen* or preteen* or pre-teen* or school-age* or teen or teens or teenager* or youth*)  Indexes=SCI-EXPANDED, SSCI, A&HCI, CPCI-S, CPCI-SSH, BKCI-S, BKCI-SSH, ESCI Timespan=All years |
| # 4 | [8,001](https://apps-webofknowledge-com.myaccess.library.utoronto.ca/summary.do?product=WOS&doc=1&qid=4&SID=5ElQLmIsRtNxI8dydUD&search_mode=CombineSearches&update_back2search_link_param=yes) | #3 OR #2 OR #1  Indexes=SCI-EXPANDED, SSCI, A&HCI, CPCI-S, CPCI-SSH, BKCI-S, BKCI-SSH, ESCI Timespan=All years |
| # 3 | [7,162](https://apps-webofknowledge-com.myaccess.library.utoronto.ca/summary.do?product=WOS&doc=1&qid=3&SID=5ElQLmIsRtNxI8dydUD&search_mode=GeneralSearch&update_back2search_link_param=yes) | TITLE: (coronavirus* or (corona W/0 virus*))  Indexes=SCI-EXPANDED, SSCI, A&HCI, CPCI-S, CPCI-SSH, BKCI-S, BKCI-SSH, ESCI Timespan=All years |
| # 2 | [523](https://apps-webofknowledge-com.myaccess.library.utoronto.ca/summary.do?product=WOS&doc=1&qid=2&SID=5ElQLmIsRtNxI8dydUD&search_mode=GeneralSearch&update_back2search_link_param=yes) | TOPIC: (coronavirus* NEAR/2 "2019") OR TOPIC: ((corona W/0 virus*) NEAR/2 "2019") OR TOPIC: (coronavirus* NEAR/2 "19") OR TOPIC: ((corona W/0 virus*) NEAR/2 "19") OR TOPIC: ((novel W/0 coronavirus*) or ("novel corona" W/0 virus*)) OR TOPIC: ("coronavirus 2" or "corona virus 2")  Indexes=SCI-EXPANDED, SSCI, A&HCI, CPCI-S, CPCI-SSH, BKCI-S, BKCI-SSH, ESCI Timespan=All years |
| # 1 | [1,234](https://apps-webofknowledge-com.myaccess.library.utoronto.ca/summary.do?product=WOS&doc=1&qid=1&SID=5ElQLmIsRtNxI8dydUD&search_mode=GeneralSearch&update_back2search_link_param=yes) | TOPIC: ("COVID-19" or COVID19) OR TOPIC: (coronavirus* and (hubei or wuhan or beijing or shanghai)) OR TOPIC: (Wuhan W/0 virus*) OR TOPIC: ("2019-nCoV") OR TOPIC: (nCoV or "n-CoV") OR TOPIC: ("HCoV-19") OR TOPIC: ("SARS-CoV-2" or "SARS-CoV2" or "SARSCoV-2" or SARSCoV2)  Indexes=SCI-EXPANDED, SSCI, A&HCI, CPCI-S, CPCI-SSH, BKCI-S, BKCI-SSH, ESCI Timespan=All years |

***************************

Update

2020 Oct 31

Ovid Multifile

Database: Ovid MEDLINE: Epub Ahead of Print, In-Process & Other Non-Indexed Citations, Ovid MEDLINE® Daily and Ovid MEDLINE® <1946-Present>, Embase Classic+Embase <1947 to 2020 October 29> , APA PsycInfo <1806 to October Week 4 2020>

Search Strategy:

--------------------------------------------------------------------------------

1	Coronavirus/ (11726)

2	Coronavirus Infections/ (38463)

3	(COVID-19 or COVID19).mp. (126503)

4	((coronavirus* or corona virus*) and (hubei or wuhan or beijing or shanghai)).mp. (7739)

5	Wuhan virus*.mp. (24)

6	2019-nCoV.mp. (2540)

7	(nCoV or n-CoV).mp. (2793)

8	HCoV-19.mp. (40)

9	(SARS-CoV-2 or SARS-CoV2 or SARSCoV-2 or SARSCoV2).mp. (43049)

10	(novel coronavirus* or novel corona virus*).mp. (12029)

11	((coronavirus* or corona virus*) adj2 "2019").mp. (74817)

12	((coronavirus* or corona virus*) adj2 "19").mp. (5177)

13	(coronavirus 2 or corona virus 2).mp. (52025)

14	(coronavirus* or corona virus*).ti. (31857)

15	or/1-14 [COVID 19] (160667)

16	Developmental Disabilities/ (46010)

17	(development* adj2 (abnormal* or atypical* or a-typical* or deficit* or delay* or deviation* or disabil* or disabled or disorder* or disturb* or handicap* or impair*)).tw,kf. (193329)

18	(global adj2 delay*).tw,kf. (4538)

19	exp Child Development Disorders, Pervasive/ (109409)

20	(autis* or asperger*).tw,kf. (173231)

21	Attention Deficit Disorder with Hyperactivity/ (73375)

22	attention deficit*.tw,kf. (102802)

23	ADHD.tw,kf. (91984)

24	hyperkinetic syndrome*.tw,kf. (1101)

25	(kanner* adj syndrome?).tw,kf. (89)

26	exp Learning Disabilities/ (88134)

27	((learning or scholastic*) adj3 (atypical* or a-typical* or deficit* or delay* or disabil* or disabled or disorder* or disturb* or dysfunction* or handicap* or impair* or retard*)).tw,kf. (102201)

28	(alexia* or alexic* or dyslexia* or dyslexic* or reading disab* or word blindness or (verbal* adj1 agnosi*)).tw,kf. (30584)

29	exp Language Development Disorders/ (11028)

30	((language or auditor* or semantic* or speech or speak* or talk* or verbal*) adj3 (atypical* or a-typical* or deficit* or delay* or disabil* or disabled or disorder* or disturb* or dysfunction* or handicap* or impair* or retard*)).tw,kf. (119545)

31	Cerebral Palsy/ (67986)

32	((cerebral adj1 palsy) or (diplegia* adj1 spastic) or (little adj1 disease)).tw,kf. (68546)

33	((brain? or central*) adj1 palsy).tw,kf. (237)

34	((brain or central* or cerebral*) adj1 (paralys* or pares#s)).tw,kf. (1773)

35	encephalopathia infantilis.tw,kf. (0)

36	exp Malformations of Cortical Development/ (19283)

37	Neural Tube Defects/ (17853)

38	(neural tube? adj2 defect*).tw,kf. (17682)

39	(acrania? or craniorachischis* or diastematomyelia* or exencephal* or iniencephal* or neurenteric cyst? or neuroenteric cyst? or spinal dysraphism* or spinal cord myelodysplasia* or (tethered adj2 cord syndrome*)).tw,kf. (9062)

40	spina? bifida*.tw,kf. (19236)

41	(cleft spine? or open spine? or rachischis* or schistorrhach* or spina? dysraphia* or status dysraphicus).tw,kf. (693)

42	Intellectual Disability/ (62896)

43	(intellectual* adj2 (atypical* or a-typical* or deficit* or delay* or deviation* or disabil* or disabled or disorder* or disturb* or dysfunction* or handicap* or impair* or retard*)).tw,kf. (72656)

44	(brain* adj2 (atypical* or a-typical* or deficit* or delay* or deviation* or disabil* or disabled or disorder* or disturb* or dysfunction* or handicap* or impair* or retard*)).tw,kf. (65161)

45	(cognitiv* adj2 (atypical* or a-typical* or deficit* or delay* or deviation* or disabil* or disabled or disorder* or disturb* or dysfunction* or handicap* or impair* or retard*)).tw,kf. (350185)

46	(cognition* adj2 (atypical* or a-typical* or deficit* or delay* or deviation* or disabil* or disabled or disorder* or disturb* or dysfunction* or handicap* or impair* or retard*)).tw,kf. (16695)

47	(mental* adj2 (atypical* or a-typical* or deficit* or delay* or deviation* or disabil* or disabled or disorder* or disturb* or dysfunction* or handicap* or impair* or retard*)).tw,kf. (335811)

48	Down Syndrome/ (68896)

49	(Down* adj2 syndrome*).tw,kf. (60709)

50	(mongolism* or mongoloid*).tw,kf. (7419)

51	("trisomy 21" or "trisomy G1" or "trisomy (G)1" or "trisomy G-1" or "trisomy GM" or "trisomy G" or "21 trisomy" or "G1 trisomy" or "G(1) trisomy" or "G-1 trisomy" or "GM trisomy" or "G trisomy").tw,kf. (15776)

52	"Chromosomes, Human, Pair 21"/ (10454)

53	(translocat* adj1 DS).tw,kf. (11)

54	Fetal Alcohol Spectrum Disorders/ (9074)

55	((f?etal or f?etus*) adj2 alcohol syndrome*).tw,kf. (6887)

56	FASD.tw,kf. (4914)

57	(alcohol-related adj3 (birth defect* or birth disorder*)).tw,kf. (302)

58	(alcohol-related adj3 (neurodevelopment* or neuro-development*) adj2 (deficit* or delay* or deviation* or disabil* or disabled or disorder* or disturb* or handicap* or impair*)).tw,kf. (368)

59	Neonatal Abstinence Syndrome/ (2325)

60	((neonat* or newborn*) adj2 (abstinen* or addiction* or withdrawal*)).tw,kf. (3835)

61	Fragile X Syndrome/ (15639)

62	"Fragile X".tw,kf. (18357)

63	(FRAXA or FRAXE or "Fra(X)").tw,kf. (1639)

64	("Mar (X)" or "Marker X" or "Martin-Bell").tw,kf. (615)

65	("X-Linked" adj2 mental retardation).tw,kf. (2832)

66	Angelman Syndrome/ (3514)

67	(angelman* or "happy puppet" or puppet child*).tw,kf. (4719)

68	exp Brain Injuries/ (285580)

69	(brain* adj3 (injur* or commotio* or concuss* or damag* or trauma*)).tw,kf. (294407)

70	concussion*.tw,kf. (25174)

71	commotio.tw,kf. (1073)

72	exp Stroke/ (379714)

73	stroke?.tw,kf. (701609)

74	((cerebrovascular or cerebro-vascular) adj2 (accident* or arrest* or failure* or injur* or insufficienc* or insult*)).tw,kf. (26138)

75	(cerebral vascular adj2 (accident* or arrest* or failure* or injur* or insufficienc* or insult*)).tw,kf. (3810)

76	isch?emic seizure*.tw,kf. (110)

77	isch?emic cerebral attack*.tw,kf. (72)

78	(apoplex* or apoplectic*).tw,kf. (8855)

79	((brain or cerebral) adj2 insult$2).tw,kf. (5568)

80	Asphyxia Neonatorum/ (11456)

81	((neonat* or newborn*) adj3 (anoxi* or asphyxi* or hypoxi* or respiratory failure*)).tw,kf. (20430)

82	Hypoxia-Ischemia, Brain/ (8846)

83	exp Infant, Premature/ (174633)

84	((preterm or pre-term or prematur* or pre-matur*) adj3 (baby or babies or infant* or neonat* or newborn*)).tw,kf. (155142)

85	exp Infant, Low Birth Weight/ (98754)

86	small for gestational age?.tw,kf. (25592)

87	(SGA or LBW or VLBW).tw,kf. (40094)

88	low* birth weight?.tw,kf. (72964)

89	exp Congenital Heart Defects/ (152311)

90	((cardiac* or cardio* or heart?) adj2 (abnormalit* or anomal* or atypical* or a-typical* or defect* or deficien* or deform* or impair* or malform*)).tw,kf. (125977)

91	(tetralog* adj2 fallot*).tw,kf. (24505)

92	((cardiac* or cardio* or heart?) adj5 (congenital* or inborn* or hereditar* or inherit*)).tw,kf. (136608)

93	Intensive Care Units, Neonatal/ and (Survivors/ or Survivorship/ or "Patient Discharge"/) (1214)

94	(neonat* adj2 (critical care or intensive care or ICU or ICUs) adj3 (discharg* or graduat* or surviv*)).tw,kf. (1654)

95	(post* adj2 (NICU or NICUs)).tw,kf. (208)

96	((NICU or NICUs) adj3 (discharg* or graduat* or surviv*)).tw,kf. (1778)

97	or/16-96 [BRAIN/HEART DISABILITIES] (3394297)

98	exp Substance-Related Disorders/ (538421)

99	(alcoholis* or alcoholic*).tw,kf. (260043)

100	((binge? or binging) adj2 (alcohol* or drink*)).tw,kf. (20125)

101	(alcohol* adj3 (abus* or addict* or dependen* or disorder? or habituat* or misus* or mis-us*)).tw,kf. (174692)

102	((narcotic* or opioid* or opiate*) adj3 (abus* or addict* or dependen* or disorder? or habituat* or misus* or mis-us* or non-medical* or nonmedical* or non-prescrib* or nonprescrib* or non-prescription* or nonprescription* or withdrawal* or (("use" or used or uses or using) adj2 (illicit* or illegal*)))).tw,kf. (60839)

103	((drug? or substance?) adj3 (abus* or addict* or dependen* or disorder? or habituat* or misus* or mis-us* or "non-medical use?" or "nonmedical use?" or "non-prescribed use?" or "nonprescribed use?" or "non-prescription use?" or "nonprescription use?" or (("use" or used or uses or using) adj2 (illicit* or illegal*)))).tw,kf. (357240)

104	(heroin* adj3 (abus* or addict* or dependen* or disorder? or habituat* or misus* or mis-us* or non-medical* or nonmedical* or non-prescrib* or nonprescrib* or non-prescription* or nonprescription* or withdrawal* or (("use" or used or uses or using) adj2 (illicit* or illegal*)))).tw,kf. (17603)

105	((hydrocodone or bekadid$2 or codinovo$2 or dico$2 or dicodid$2 or dihydrocodeinone$2 or hycodan$2 or hycon$2 or hydrocodeinonebitartrate$2 or hydrocodon$2 or hydrocon$2 or hydrocodonum$2 or robidone$2) adj3 (abus* or addict* or dependen* or disorder? or habituat* or misus* or mis-us* or non-medical* or nonmedical* or non-prescrib* or nonprescrib* or non-prescription* or nonprescription* or withdrawal* or (("use" or used or uses or using) adj2 (illicit* or illegal*)))).tw,kf. (158)

106	((fentanyl or alfentanil$2 or alfenta$2 or alfentanyl$2 or beta hydroxymefentanyl or brifentanil$2 or carfentanil$2 or duragesic$2 or fanaxal$2 or fentanest$2 or fentora$2 or hypnorm$2 or limifen$2 or lofentanil$2 or mefentanyl$2 or mirfentanil$2 or ocfentanil$2 or phentanyl$2 or R-39209 or R-4263 or rapifen$2 or remifentanil$2 or sublimaze$2 or sufenta$2 or sufentanil$2 or sulfentanyl$2 or trefentanil$2) adj3 (abus* or addict* or dependen* or disorder? or habituat* or misus* or mis-us* or non-medical* or nonmedical* or non-prescrib* or nonprescrib* or non-prescription* or nonprescription* or withdrawal* or (("use" or used or uses or using) adj2 (illicit* or illegal*)))).tw,kf. (905)

107	((morphine or anpec$2 or duramorph$2 or epimorph$2 or miro$2 or morfin$2 or morfine$2 or morphin$2 or morphinium$2 or morphium$2 or MS contin or morphia$2 or opso$2 or oramorph$2 or SDZ 202-250 or SDZ202-250 or skenan$2 or transmorphine$2 or trans-morphine$2) adj3 (abus* or addict* or dependen* or disorder? or habituat* or misus* or mis-us* or non-medical* or nonmedical* or non-prescrib* or nonprescrib* or non-prescription* or nonprescription* or withdrawal* or (("use" or used or uses or using) adj2 (illicit* or illegal*)))).tw,kf. (12837)

108	((oxycodone or bionine$2 or bionone$2 or bolodorm$2 or broncodal$2 or bucodal$2 or cafacodal$2 or cardanon$2 or codenon$2 or codix 5 or "col 003" or col003 or DETERx$2 or dihydrohydroxycodeinone or dihydrohydroxydodeinone or dihydrone$2 or dinarkon$2 or endone$2 or eubine$2 or eucodal$2 or eucodale$2 or eucodalum$2 or eudin$2 or eukdin$2 or eukodal$2 or eumorphal$2 or eurodamine$2 or eutagen$2 or hydrocodal$2 or hydroxycodeinoma$2 or ludonal$2 or m-oxy or medicodal$2 or narcobasina$2 or narcobasine$2 or narcosin$2 or nargenol$2 or narodal$2 or nsc 19043 or nucodan$2 or opton$2 or ossicodone$2 or oxanest$2 or oxaydo$2 or oxecta$2 or oxicone$2 or oxicontin$2 or oxiconum$2 or oxikon$2 or oxy ir or oxycod$2 or oxycodeinon$2 or oxycodeinonhydrochloride or oxycodone hydrochloride or oxycodonhydrochlorid or oxycodyl$2 or oxycone$2 or oxycontin$2 or oxydose$2 or oxyfast$2 or oxygesic$2 or oxyir$2 or oxykon$2 or oxynorm$2 or pancodine$2 or pavinal$2 or percolone$2 or pronarcin$2 or remoxy$2 or roxicodone$2 or roxycodone$2 or sinthiodal$2 or stupenal$2 or supeudol$2 or tebodal$2 or tekodin$2 or thecodin$2 or theocodin$2 or xtampa$2 or xtampza$2) adj3 (abus* or addict* or dependen* or disorder? or habituat* or misus* or mis-us* or non-medical* or nonmedical* or non-prescrib* or nonprescrib* or non-prescription* or nonprescription* or withdrawal* or (("use" or used or uses or using) adj2 (illicit* or illegal*)))).tw,kf. (736)

109	or/98-108 [SUBSTANCE/ALCOHOL DISORDERS] (1017135)

110	exp Infant/ (2323956)

111	exp Child/ (5000102)

112	Adolescent/ (3734371)

113	(baby or babies or infant? or infanc* or neonat* or newborn* or preschool* or pre-school* or toddler?).tw,kf. (1974884)

114	(adolescen* or child* or pre-adolescen* or preteen* or pre-teen* or school-age* or teen or teens or teenager? or youth*).tw,kf. (4771870)

115	exp Pediatrics/ (210118)

116	p?ediatric*.tw,kf. (986896)

117	or/110-116 [INFANTS/CHILDREN/ADOLESCENTS] (10033552)

118	97 and 117 [BRAIN DISABILITIES - CHILDREN/ADOLESCENTS/YOUNG ADULTS] (1274649)

119	15 and 118 [COVID-19 - BRAIN DISABILITIES - CHILDREN/ADOLESCENTS/YOUNG ADULTS] (785)

120	Prenatal Exposure Delayed Effects/ (53524)

121	(prenatal* or pre-natal* or preterm* or antenatal* or ante-natal* or antepartum* or ante-partum*).tw,kf. (525883)

122	((before or prior) adj3 (birth* or childbirth* or child birth*)).tw,kf. (25864)

123	(baby or babies or infant? or infanc* or neonat* or newborn*).tw,kf. (1850894)

124	or/120-123 [PRENATAL EXPOSURE/NEWBORNS] (2167152)

125	109 and 124 [SUBSTANCE ABUSE - PRENATAL EXPOSURE/NEWBORNS] (28800)

126	15 and 125 [COVID-19 - SUBSTANCE ABUSE - PRENATAL EXPOSURE/NEWBORNS] (1)

127	119 or 126 [ALL CONDITIONS OF INTEREST] (785)

128	exp Animals/ not Humans/ (18591781)

129	127 not 128 [ANIMAL-ONLY REMOVED] (392)

130	limit 129 to yr="2019-current" (364)

131	(2020041* or 2020042* or "20200430" or 202005* or 202006* or 202007* or 202008* or 202009* or 202010*).dt. (850688)

132	130 and 131 [UPDATE PERIOD] (296)

133	132 use ppez [MEDLINE RECORDS] (296)

134	Coronavirinae/ (4154)

135	Coronavirus infection/ (49107)

136	(COVID-19 or COVID19).mp. (126503)

137	((coronavirus* or corona virus*) and (hubei or wuhan or beijing or shanghai)).mp. (7739)

138	Wuhan virus*.mp. (24)

139	2019-nCoV.mp. (2540)

140	(nCoV or n-CoV).mp. (2793)

141	HCoV-19.mp. (40)

142	(SARS-CoV-2 or SARS-CoV2 or SARSCoV-2 or SARSCoV2).mp. (43049)

143	(novel coronavirus* or novel corona virus*).mp. (12029)

144	((coronavirus* or corona virus*) adj2 "2019").mp. (74817)

145	((coronavirus* or corona virus*) adj2 "19").mp. (5177)

146	(coronavirus 2 or corona virus 2).mp. (52025)

147	(coronavirus* or corona virus*).ti. (31857)

148	or/134-147 [COVID 19] (159105)

149	exp developmental disorder/ (47316)

150	(development* adj2 (abnormal* or atypical* or a-typical* or deficit* or delay* or deviation* or disabil* or disabled or disorder* or disturb* or handicap* or impair*)).tw,kw. (194866)

151	(global adj2 delay*).tw,kw. (4532)

152	exp autism/ (138455)

153	(autis* or asperger*).tw,kw. (174912)

154	attention deficit disorder/ (94217)

155	attention deficit*.tw,kw. (104003)

156	ADHD.tw,kw. (93039)

157	hyperkinetic syndrome*.tw,kw. (1117)

158	(kanner* adj syndrome?).tw,kw. (90)

159	exp learning disorder/ (72466)

160	((learning or scholastic*) adj3 (atypical* or a-typical* or deficit* or delay* or disabil* or disabled or disorder* or disturb* or dysfunction* or handicap* or impair* or retard*)).tw,kw. (102558)

161	(alexia* or alexic* or dyslexia* or dyslexic* or reading disab* or word blindness or (verbal* adj1 agnosi*)).tw,kw. (31041)

162	exp developmental language disorder/ (11028)

163	((language or auditor* or semantic* or speech or speak* or talk* or verbal*) adj3 (atypical* or a-typical* or deficit* or delay* or disabil* or disabled or disorder* or disturb* or dysfunction* or handicap* or impair* or retard*)).tw,kw. (118966)

164	cerebral palsy/ (67986)

165	((cerebral adj1 palsy) or (diplegia* adj1 spastic) or (little adj1 disease)).tw,kw. (68484)

166	((brain? or central*) adj1 palsy).tw,kw. (238)

167	((brain or central* or cerebral*) adj1 (paralys* or pares#s)).tw,kw. (1870)

168	encephalopathia infantilis.tw,kw. (0)

169	exp cortical dysplasia/ (19283)

170	exp neural tube defect/ (62884)

171	(neural tube? adj2 defect*).tw,kw. (18055)

172	(acrania? or craniorachischis* or diastematomyelia* or exencephal* or iniencephal* or neurenteric cyst? or neuroenteric cyst? or spinal dysraphism* or spinal cord myelodysplasia* or (tethered adj2 cord syndrome*)).tw,kw. (9552)

173	spina? bifida*.tw,kw. (19985)

174	(cleft spine? or open spine? or rachischis* or schistorrhach* or spina? dysraphia* or status dysraphicus).tw,kw. (707)

175	exp intellectual impairment/ (551411)

176	(intellectual* adj2 (atypical* or a-typical* or deficit* or delay* or deviation* or disabil* or disabled or disorder* or disturb* or dysfunction* or handicap* or impair* or retard*)).tw,kw. (73165)

177	(brain* adj2 (atypical* or a-typical* or deficit* or delay* or deviation* or disabil* or disabled or disorder* or disturb* or dysfunction* or handicap* or impair* or retard*)).tw,kw. (65706)

178	(cognitiv* adj2 (atypical* or a-typical* or deficit* or delay* or deviation* or disabil* or disabled or disorder* or disturb* or dysfunction* or handicap* or impair* or retard*)).tw,kw. (352637)

179	(cognition* adj2 (atypical* or a-typical* or deficit* or delay* or deviation* or disabil* or disabled or disorder* or disturb* or dysfunction* or handicap* or impair* or retard*)).tw,kw. (17472)

180	(mental* adj2 (atypical* or a-typical* or deficit* or delay* or deviation* or disabil* or disabled or disorder* or disturb* or dysfunction* or handicap* or impair* or retard*)).tw,kw. (331515)

181	Down syndrome/ (68896)

182	(Down* adj2 syndrome*).tw,kw. (61259)

183	(mongolism* or mongoloid*).tw,kw. (7532)

184	trisomy 21/ (39549)

185	("trisomy 21" or "trisomy G1" or "trisomy (G)1" or "trisomy G-1" or "trisomy GM" or "trisomy G" or "21 trisomy" or "G1 trisomy" or "G(1) trisomy" or "G-1 trisomy" or "GM trisomy" or "G trisomy").tw,kw. (16349)

186	chromosome 21/ (10454)

187	(translocat* adj1 DS).tw,kw. (11)

188	fetal alcohol syndrome/ (12900)

189	((f?etal or f?etus*) adj2 alcohol syndrome*).tw,kw. (7441)

190	FASD.tw,kw. (5012)

191	(alcohol-related adj3 (birth defect* or birth disorder*)).tw,kw. (308)

192	(alcohol-related adj3 (neurodevelopment* or neuro-development*) adj2 (deficit* or delay* or deviation* or disabil* or disabled or disorder* or disturb* or handicap* or impair*)).tw,kw. (388)

193	neonatal abstinence syndrome/ (2325)

194	((neonat* or newborn*) adj2 (abstinen* or addiction* or withdrawal*)).tw,kw. (3895)

195	fragile X syndrome/ (15639)

196	"Fragile X".tw,kw. (18535)

197	(FRAXA or FRAXE or "Fra(X)").tw,kw. (1663)

198	("Mar (X)" or "Marker X" or "Martin-Bell").tw,kw. (630)

199	("X-Linked" adj2 mental retardation).tw,kw. (2982)

200	happy puppet syndrome/ (4021)

201	(angelman* or "happy puppet" or puppet child*).tw,kw. (4797)

202	exp brain injury/ (285196)

203	(brain* adj3 (injur* or commotio* or concuss* or damag* or trauma*)).tw,kw. (295289)

204	concussion*.tw,kw. (26300)

205	commotio.tw,kw. (1099)

206	exp cerebrovascular accident/ (379714)

207	stroke?.tw,kw. (711941)

208	((cerebrovascular or cerebro-vascular) adj2 (accident* or arrest* or failure* or injur* or insufficienc* or insult*)).tw,kw. (27361)

209	(cerebral vascular adj2 (accident* or arrest* or failure* or injur* or insufficienc* or insult*)).tw,kw. (3866)

210	isch?emic seizure*.tw,kw. (111)

211	isch?emic cerebral attack*.tw,kw. (72)

212	(apoplex* or apoplectic*).tw,kw. (8957)

213	((brain or cerebral) adj2 insult$2).tw,kw. (5589)

214	newborn hypoxia/ (6901)

215	((neonat* or newborn*) adj3 (anoxi* or asphyxi* or hypoxi* or respiratory failure*)).tw,kw. (20220)

216	hypoxic ischemic encephalopathy/ (13532)

217	Prematurity/ (118110)

218	((preterm or pre-term or prematur* or pre-matur*) adj3 (baby or babies or infant* or neonat* or newborn*)).tw,kw. (155041)

219	exp low birth weight/ (101984)

220	small for gestational age?.tw,kw. (26035)

221	(SGA or LBW or VLBW).tw,kw. (40359)

222	low* birth weight?.tw,kw. (73931)

223	exp congenital heart malformation/ (155668)

224	((cardiac* or cardio* or heart?) adj2 (abnormalit* or anomal* or atypical* or a-typical* or defect* or deficien* or deform* or impair* or malform*)).tw,kw. (123344)

225	(tetralog* adj2 fallot*).tw,kw. (24360)

226	((cardiac* or cardio* or heart?) adj5 (congenital* or inborn* or hereditar* or inherit*)).tw,kw. (138650)

227	neonatal intensive care unit/ and (exp survivor/ or survivorship/ or hospital discharge.mp.) [mp=ti, ab, ot, nm, hw, fx, kf, ox, px, rx, ui, sy, tn, dm, mf, dv, kw, dq, tc, id, tm, mh] (1039)

228	(neonat* adj2 (critical care or intensive care or ICU or ICUs) adj3 (discharg* or graduat* or surviv*)).tw,kw. (1657)

229	(post* adj2 (NICU or NICUs)).tw,kw. (219)

230	((NICU or NICUs) adj3 (discharg* or graduat* or surviv*)).tw,kw. (1785)

231	or/149-229 [BRAIN/HEART DISABILITIES] (3707184)

232	exp drug dependence/ (538421)

233	(alcoholis* or alcoholic*).tw,kw. (263269)

234	((binge? or binging) adj2 (alcohol* or drink*)).tw,kw. (20259)

235	(alcohol* adj3 (abus* or addict* or dependen* or disorder? or habituat* or misus* or mis-us*)).tw,kw. (175979)

236	((narcotic* or opioid* or opiate*) adj3 (abus* or addict* or dependen* or disorder? or habituat* or misus* or mis-us* or non-medical* or nonmedical* or non-prescrib* or nonprescrib* or non-prescription* or nonprescription* or withdrawal* or (("use" or used or uses or using) adj2 (illicit* or illegal*)))).tw,kw. (61703)

237	((drug? or substance?) adj3 (abus* or addict* or dependen* or disorder? or habituat* or misus* or mis-us* or "non-medical use?" or "nonmedical use?" or "non-prescribed use?" or "nonprescribed use?" or "non-prescription use?" or "nonprescription use?" or (("use" or used or uses or using) adj2 (illicit* or illegal*)))).tw,kw. (360765)

238	(heroin* adj3 (abus* or addict* or dependen* or disorder? or habituat* or misus* or mis-us* or non-medical* or nonmedical* or non-prescrib* or nonprescrib* or non-prescription* or nonprescription* or withdrawal* or (("use" or used or uses or using) adj2 (illicit* or illegal*)))).tw,kw. (17827)

239	((hydrocodone or bekadid$2 or codinovo$2 or dico$2 or dicodid$2 or dihydrocodeinone$2 or hycodan$2 or hycon$2 or hydrocodeinonebitartrate$2 or hydrocodon$2 or hydrocon$2 or hydrocodonum$2 or robidone$2) adj3 (abus* or addict* or dependen* or disorder? or habituat* or misus* or mis-us* or non-medical* or nonmedical* or non-prescrib* or nonprescrib* or non-prescription* or nonprescription* or withdrawal* or (("use" or used or uses or using) adj2 (illicit* or illegal*)))).tw,kw. (171)

240	((fentanyl or alfentanil$2 or alfenta$2 or alfentanyl$2 or beta hydroxymefentanyl or brifentanil$2 or carfentanil$2 or duragesic$2 or fanaxal$2 or fentanest$2 or fentora$2 or hypnorm$2 or limifen$2 or lofentanil$2 or mefentanyl$2 or mirfentanil$2 or ocfentanil$2 or phentanyl$2 or R-39209 or R-4263 or rapifen$2 or remifentanil$2 or sublimaze$2 or sufenta$2 or sufentanil$2 or sulfentanyl$2 or trefentanil$2) adj3 (abus* or addict* or dependen* or disorder? or habituat* or misus* or mis-us* or non-medical* or nonmedical* or non-prescrib* or nonprescrib* or non-prescription* or nonprescription* or withdrawal* or (("use" or used or uses or using) adj2 (illicit* or illegal*)))).tw,kw. (930)

241	((morphine or anpec$2 or duramorph$2 or epimorph$2 or miro$2 or morfin$2 or morfine$2 or morphin$2 or morphinium$2 or morphium$2 or MS contin or morphia$2 or opso$2 or oramorph$2 or SDZ 202-250 or SDZ202-250 or skenan$2 or transmorphine$2 or trans-morphine$2) adj3 (abus* or addict* or dependen* or disorder? or habituat* or misus* or mis-us* or non-medical* or nonmedical* or non-prescrib* or nonprescrib* or non-prescription* or nonprescription* or withdrawal* or (("use" or used or uses or using) adj2 (illicit* or illegal*)))).tw,kw. (12879)

242	((oxycodone or bionine$2 or bionone$2 or bolodorm$2 or broncodal$2 or bucodal$2 or cafacodal$2 or cardanon$2 or codenon$2 or codix 5 or "col 003" or col003 or DETERx$2 or dihydrohydroxycodeinone or dihydrohydroxydodeinone or dihydrone$2 or dinarkon$2 or endone$2 or eubine$2 or eucodal$2 or eucodale$2 or eucodalum$2 or eudin$2 or eukdin$2 or eukodal$2 or eumorphal$2 or eurodamine$2 or eutagen$2 or hydrocodal$2 or hydroxycodeinoma$2 or ludonal$2 or m-oxy or medicodal$2 or narcobasina$2 or narcobasine$2 or narcosin$2 or nargenol$2 or narodal$2 or nsc 19043 or nucodan$2 or opton$2 or ossicodone$2 or oxanest$2 or oxaydo$2 or oxecta$2 or oxicone$2 or oxicontin$2 or oxiconum$2 or oxikon$2 or oxy ir or oxycod$2 or oxycodeinon$2 or oxycodeinonhydrochloride or oxycodone hydrochloride or oxycodonhydrochlorid or oxycodyl$2 or oxycone$2 or oxycontin$2 or oxydose$2 or oxyfast$2 or oxygesic$2 or oxyir$2 or oxykon$2 or oxynorm$2 or pancodine$2 or pavinal$2 or percolone$2 or pronarcin$2 or remoxy$2 or roxicodone$2 or roxycodone$2 or sinthiodal$2 or stupenal$2 or supeudol$2 or tebodal$2 or tekodin$2 or thecodin$2 or theocodin$2 or xtampa$2 or xtampza$2) adj3 (abus* or addict* or dependen* or disorder? or habituat* or misus* or mis-us* or non-medical* or nonmedical* or non-prescrib* or nonprescrib* or non-prescription* or nonprescription* or withdrawal* or (("use" or used or uses or using) adj2 (illicit* or illegal*)))).tw,kw. (770)

243	or/232-242 [SUBSTANCE/ALCOHOL DISORDERS] (1021957)

244	exp child/ (5000102)

245	exp adolescent/ (3734583)

246	juvenile/ (52115)

247	(baby or babies or infant? or infanc* or neonat* or newborn* or preschool* or pre-school* or toddler?).tw,kw. (1956842)

248	(adolescen* or child* or pre-adolescen* or preteen* or pre-teen* or school-age* or teen or teens or teenager? or youth*).tw,kw. (4779185)

249	exp pediatrics/ (210118)

250	p?ediatric*.tw,kw. (1014514)

251	or/244-250 [INFANTS/CHILDREN/ADOLESCENTS] (9875135)

252	231 and 251 [BRAIN DISABILITIES - CHILDREN/ADOLESCENTS/YOUNG ADULTS] (1271361)

253	148 and 252 [COVID-19 - BRAIN DISABILITIES - CHILDREN/ADOLESCENTS/YOUNG ADULTS] (790)

254	prenatal exposure/ (61304)

255	(prenatal* or pre-natal* or preterm* or antenatal* or ante-natal* or antepartum* or ante-partum*).tw,kw. (533139)

256	((before or prior) adj3 (birth* or childbirth* or child birth*)).tw,kw. (25871)

257	(baby or babies or infant? or infanc* or neonat* or newborn*).tw,kw. (1831477)

258	or/254-257 [PRENATAL EXPOSURE/NEWBORNS] (2152814)

259	243 and 258 [SUBSTANCE ABUSE - PRENATAL EXPOSURE/NEWBORNS] (29286)

260	148 and 259 [COVID-19 - SUBSTANCE ABUSE - PRENATAL EXPOSURE/NEWBORNS] (1)

261	253 or 260 [ALL CONDITIONS OF INTEREST] (790)

262	exp animal/ or exp animal experimentation/ or exp animal model/ or exp animal experiment/ or nonhuman/ or exp vertebrate/ (54231358)

263	exp human/ or exp human experimentation/ or exp human experiment/ (41754484)

264	262 not 263 (12478679)

265	261 not 264 [ANIMAL-ONLY REMOVED] (786)

266	limit 265 to yr="2019-current" (743)

267	(2020041* or 2020042* or "20200430" or 202005* or 202006* or 202007* or 202008* or 202009* or 202010*).dc. (1339945)

268	266 and 267 [UPDATE PERIOD] (414)

269	268 use emczd [EMBASE RECORDS - UPDATE PERIOD] (414)

270	(COVID-19 or COVID19).mp. (126503)

271	((coronavirus* or corona virus*) and (hubei or wuhan or beijing or shanghai)).mp. (7739)

272	Wuhan virus*.mp. (24)

273	2019-nCoV.mp. (2540)

274	(nCoV or n-CoV).mp. (2793)

275	HCoV-19.mp. (40)

276	(SARS-CoV-2 or SARS-CoV2 or SARSCoV-2 or SARSCoV2).mp. (43049)

277	(novel coronavirus* or novel corona virus*).mp. (12029)

278	((coronavirus* or corona virus*) adj2 "2019").mp. (74817)

279	((coronavirus* or corona virus*) adj2 "19").mp. (5177)

280	(coronavirus 2 or corona virus 2).mp. (52025)

281	(coronavirus* or corona virus*).ti. (31857)

282	or/270-281 [COVID 19] (153877)

283	(baby or babies or infant? or infanc* or neonat* or newborn* or preschool* or pre-school* or toddler?).tw,id. (1924606)

284	(adolescen* or child* or pre-adolescen* or preteen* or pre-teen* or school-age* or teen or teens or teenager? or youth*).tw,id. (4717264)

285	exp Pediatrics/ (210118)

286	p?ediatric*.tw,id. (970266)

287	limit 282 to (100 childhood <birth to age 12 yrs> or 120 neonatal <birth to age 1 mo> or 140 infancy <2 to 23 mo> or 160 preschool age <age 2 to 5 yrs> or 180 school age <age 6 to 12 yrs> or 200 adolescence <age 13 to 17 yrs>) [Limit not valid in Ovid MEDLINE(R),Ovid MEDLINE(R) Daily Update,Ovid MEDLINE(R) In-Process,Ovid MEDLINE(R) Publisher,Embase; records were retained] (151769)

288	283 or 284 or 285 or 286 (6408538)

289	282 and 288 (10103)

290	287 or 289 [COVID-19 - INFANT/CHILD/ADOLESCENT POPULATION] (152014)

291	limit 290 to yr="2019-current" (139236)

292	(202004* or 202005* or 202006* or 202007* or 202008* or 202009* or 202010*).up. (3809199)

293	291 and 292 [UPDATE PERIOD] (69839)

294	293 use ppez,emczd (69519)

295	293 not 294 [PSYCINFO RECORDS] (320)

296	133 or 269 or 295 [ALL DATABASES] (1030)

297	remove duplicates from 296 (777)

298	297 use ppez [MEDLINE UNIQUE RECORDS] (289)

299	297 use emczd [EMBASE UNIQUE RECORDS] (197)

300	297 not (298 or 299) [PSYCINFO UNIQUE RECORDS] (291)

***************************

CINAHL

| # | Query | Limiters/Expanders | Last Run Via | Results |
| --- | --- | --- | --- | --- |
| S89 | S87 AND S88 | Search modes - Boolean/Phrase | Interface - EBSCOhost Research Databases  Search Screen - Advanced Search  Database - CINAHL Plus with Full Text | 64 |
| S88 | EM 20200410-20201231 | Search modes - Boolean/Phrase | Interface - EBSCOhost Research Databases  Search Screen - Advanced Search  Database - CINAHL Plus with Full Text | 201,056 |
| S87 | S80 OR S85 | Limiters - Published Date: 20190101-20201231  Search modes - Boolean/Phrase | Interface - EBSCOhost Research Databases  Search Screen - Advanced Search  Database - CINAHL Plus with Full Text | 118 |
| S86 | S80 OR S85 | Search modes - Boolean/Phrase | Interface - EBSCOhost Research Databases  Search Screen - Advanced Search  Database - CINAHL Plus with Full Text | 133 |
| S85 | S9 AND S84 | Search modes - Boolean/Phrase | Interface - EBSCOhost Research Databases  Search Screen - Advanced Search  Database - CINAHL Plus with Full Text | 2 |
| S84 | S72 AND S83 | Search modes - Boolean/Phrase | Interface - EBSCOhost Research Databases  Search Screen - Advanced Search  Database - CINAHL Plus with Full Text | 5,495 |
| S83 | S81 OR S82 | Search modes - Boolean/Phrase | Interface - EBSCOhost Research Databases  Search Screen - Advanced Search  Database - CINAHL Plus with Full Text | 194,187 |
| S82 | TI ( (before or prior) N3 (birth* or childbirth* or (child N0 birth*)) ) OR AB ( (before or prior) N3 (birth* or childbirth* or (child N0 birth*)) ) OR TI ( baby or babies or infant# or infanc* or neonat* or newborn* ) OR AB ( baby or babies or infant# or infanc* or neonat* or newborn* ) | Search modes - Boolean/Phrase | Interface - EBSCOhost Research Databases  Search Screen - Advanced Search  Database - CINAHL Plus with Full Text | 190,280 |
| S81 | (MH "Prenatal Exposure Delayed Effects") | Search modes - Boolean/Phrase | Interface - EBSCOhost Research Databases  Search Screen - Advanced Search  Database - CINAHL Plus with Full Text | 5,532 |
| S80 | S9 AND S79 | Search modes - Boolean/Phrase | Interface - EBSCOhost Research Databases  Search Screen - Advanced Search  Database - CINAHL Plus with Full Text | 131 |
| S79 | S62 AND S78 | Search modes - Boolean/Phrase | Interface - EBSCOhost Research Databases  Search Screen - Advanced Search  Database - CINAHL Plus with Full Text | 180,358 |
| S78 | S73 OR S74 OR S75 OR S76 OR S77 | Search modes - Boolean/Phrase | Interface - EBSCOhost Research Databases  Search Screen - Advanced Search  Database - CINAHL Plus with Full Text | 1,245,654 |
| S77 | TI p#ediatric* OR AB p#ediatric* | Search modes - Boolean/Phrase | Interface - EBSCOhost Research Databases  Search Screen - Advanced Search  Database - CINAHL Plus with Full Text | 139,118 |
| S76 | (MH "Pediatrics+") | Search modes - Boolean/Phrase | Interface - EBSCOhost Research Databases  Search Screen - Advanced Search  Database - CINAHL Plus with Full Text | 21,413 |
| S75 | TI ( adolescen* or child* or pre-adolescen* or preteen* or pre-teen* or school-age* or teen or teens or teenager# or youth* ) OR AB ( adolescen* or child* or pre-adolescen* or preteen* or pre-teen* or school-age* or teen or teens or teenager# or youth* ) | Search modes - Boolean/Phrase | Interface - EBSCOhost Research Databases  Search Screen - Advanced Search  Database - CINAHL Plus with Full Text | 611,289 |
| S74 | TI ( baby or babies or infant# or infanc* or neonat* or newborn* or preschool* or pre-school* or toddler# ) OR AB ( baby or babies or infant# or infanc* or neonat* or newborn* or preschool* or pre-school* or toddler# ) | Search modes - Boolean/Phrase | Interface - EBSCOhost Research Databases  Search Screen - Advanced Search  Database - CINAHL Plus with Full Text | 205,298 |
| S73 | (MH "Child+") OR (MH "Minors (Legal)") OR (MH "Adolescence") | Search modes - Boolean/Phrase | Interface - EBSCOhost Research Databases  Search Screen - Advanced Search  Database - CINAHL Plus with Full Text | 990,585 |
| S72 | S63 OR S64 OR S65 OR S66 OR S67 OR S68 OR S69 OR S70 OR S71 | Search modes - Boolean/Phrase | Interface - EBSCOhost Research Databases  Search Screen - Advanced Search  Database - CINAHL Plus with Full Text | 198,740 |
| S71 | TI ( (oxycodone or bionine or bionone or bolodorm or broncodal or bucodal or cafacodal or cardanon or codenon or codix 5 or "col 003" or col003 or DETERx or dihydrohydroxycodeinone or dihydrohydroxydodeinone or dihydrone or dinarkon or endone or eubine or eucodal or eucodale or eucodalum or eudin or eukdin or eukodal or eumorphal or eurodamine or eutagen or hydrocodal or hydroxycodeinoma or ludonal or m-oxy or medicodal or narcobasina or narcobasine or narcosin or nargenol or narodal or nsc 19043 or nucodan or opton or ossicodone or oxanest or oxaydo or oxecta or oxicone or oxicontin or oxiconum or oxikon or oxy ir or oxycod or oxycodeinon or oxycodeinonhydrochloride or oxycodone hydrochloride or oxycodonhydrochlorid or oxycodyl or oxycone or oxycontin or oxydose or oxyfast or oxygesic or oxyir or oxykon or oxynorm or pancodine or pavinal or percolone or pronarcin or remoxy or roxicodone or roxycodone or sinthiodal or stupenal or supeudol or tebodal or tekodin or thecodin or theocodin or xtampa or xtampza) N3 (abus* or addict* or dependen* or disorder# or habituat* or misus* or (mis N0 us*) or (non N0 medical*) or nonmedical* or (non N0 prescrib*) or nonprescrib* or (non N0 prescription*) or nonprescription* or withdrawal* or (("use" or used or uses or using) N2 (illicit* or illegal*))) ) OR AB ( (oxycodone or bionine or bionone or bolodorm or broncodal or bucodal or cafacodal or cardanon or codenon or codix 5 or "col 003" or col003 or DETERx or dihydrohydroxycodeinone or dihydrohydroxydodeinone or dihydrone or dinarkon or endone or eubine or eucodal or eucodale or eucodalum or eudin or eukdin or eukodal or eumorphal or eurodamine or eutagen or hydrocodal or hydroxycodeinoma or ludonal or m-oxy or medicodal or narcobasina or narcobasine or narcosin or nargenol or narodal or nsc 19043 or nucodan or opton or ossicodone or oxanest or oxaydo or oxecta or oxicone or oxicontin or oxiconum or oxikon or oxy ir or oxycod or oxycodeinon or oxycodeinonhydrochloride or oxycodone hydrochloride or oxycodonhydrochlorid or oxycodyl or oxycone or oxycontin or oxydose or oxyfast or oxygesic or oxyir or oxykon or oxynorm or pancodine or pavinal or percolone or pronarcin or remoxy or roxicodone or roxycodone or sinthiodal or stupenal or supeudol or tebodal or tekodin or thecodin or theocodin or xtampa or xtampza) N3 (abus* or addict* or dependen* or disorder# or habituat* or misus* or (mis N0 us*) or (non N0 medical*) or nonmedical* or (non N0 prescrib*) or nonprescrib* or (non N0 prescription*) or nonprescription* or withdrawal* or (("use" or used or uses or using) N2 (illicit* or illegal*))) ) | Search modes - Boolean/Phrase | Interface - EBSCOhost Research Databases  Search Screen - Advanced Search  Database - CINAHL Plus with Full Text | 170 |
| S70 | TI ( (morphine or anpec or duramorph or epimorph or miro or morfin or morfine or morphin or morphinium or morphium or MS contin or morphia or opso or oramorph or SDZ 202-250 or SDZ202-250 or skenan or transmorphine or trans-morphine) (abus* or addict* or dependen* or disorder# or habituat* or misus* or (mis N0 us*) or (non N0 medical*) or nonmedical* or (non N0 prescrib*) or nonprescrib* or (non N0 prescription*) or nonprescription* or withdrawal* or (("use" or used or uses or using) N2 (illicit* or illegal*))) ) OR AB ( (morphine or anpec or duramorph or epimorph or miro or morfin or morfine or morphin or morphinium or morphium or MS contin or morphia or opso or oramorph or SDZ 202-250 or SDZ202-250 or skenan or transmorphine or trans-morphine) (abus* or addict* or dependen* or disorder# or habituat* or misus* or (mis N0 us*) or (non N0 medical*) or nonmedical* or (non N0 prescrib*) or nonprescrib* or (non N0 prescription*) or nonprescription* or withdrawal* or (("use" or used or uses or using) N2 (illicit* or illegal*))) ) | Search modes - Boolean/Phrase | Interface - EBSCOhost Research Databases  Search Screen - Advanced Search  Database - CINAHL Plus with Full Text | 157 |
| S69 | TI ( (fentanyl or alfentanil or alfenta or alfentanyl or beta hydroxymefentanyl or brifentanil or carfentanil or duragesic or fanaxal or fentanest or fentora or hypnorm or limifen or lofentanil or mefentanyl or mirfentanil or ocfentanil or phentanyl or R-39209 or R-4263 or rapifen or remifentanil or sublimaze or sufenta or sufentanil or sulfentanyl or trefentanil) N3 (abus* or addict* or dependen* or disorder# or habituat* or misus* or (mis N0 us*) or (non N0 medical*) or nonmedical* or (non N0 prescrib*) or nonprescrib* or (non N0 prescription*) or nonprescription* or withdrawal* or (("use" or used or uses or using) N2 (illicit* or illegal*))) ) OR AB ( (fentanyl or alfentanil or alfenta or alfentanyl or beta hydroxymefentanyl or brifentanil or carfentanil or duragesic or fanaxal or fentanest or fentora or hypnorm or limifen or lofentanil or mefentanyl or mirfentanil or ocfentanil or phentanyl or R-39209 or R-4263 or rapifen or remifentanil or sublimaze or sufenta or sufentanil or sulfentanyl or trefentanil) N3 (abus* or addict* or dependen* or disorder# or habituat* or misus* or (mis N0 us*) or (non N0 medical*) or nonmedical* or (non N0 prescrib*) or nonprescrib* or (non N0 prescription*) or nonprescription* or withdrawal* or (("use" or used or uses or using) N2 (illicit* or illegal*))) ) | Search modes - Boolean/Phrase | Interface - EBSCOhost Research Databases  Search Screen - Advanced Search  Database - CINAHL Plus with Full Text | 141 |
| S68 | TI ( (hydrocodone or bekadid or codinovo or dico or dicodid or dihydrocodeinone or hycodan or hycon or hydrocodeinonebitartrate or hydrocodon or hydrocon or hydrocodonum or robidone) N3 (abus* or addict* or dependen* or disorder# or habituat* or misus* or (mis N0 us*) or (non N0 medical*) or nonmedical* or (non N0 prescrib*) or nonprescrib* or (non N0 prescription*) or nonprescription* or withdrawal* or (("use" or used or uses or using) N2 (illicit* or illegal*))) ) OR AB ( (hydrocodone or bekadid or codinovo or dico or dicodid or dihydrocodeinone or hycodan or hycon or hydrocodeinonebitartrate or hydrocodon or hydrocon or hydrocodonum or robidone) N3 (abus* or addict* or dependen* or disorder# or habituat* or misus* or (mis N0 us*) or (non N0 medical*) or nonmedical* or (non N0 prescrib*) or nonprescrib* or (non N0 prescription*) or nonprescription* or withdrawal* or (("use" or used or uses or using) N2 (illicit* or illegal*))) ) | Search modes - Boolean/Phrase | Interface - EBSCOhost Research Databases  Search Screen - Advanced Search  Database - CINAHL Plus with Full Text | 43 |
| S67 | TI ( heroin* N3 (abus* or addict* or dependen* or disorder# or habituat* or misus* or (mis N0 us*) or (non N0 medical*) or nonmedical* or (non N0 prescrib*) or nonprescrib* or (non N0 prescription*) or nonprescription* or withdrawal* or (("use" or used or uses or using) N2 (illicit* or illegal*))) ) OR AB ( heroin* N3 (abus* or addict* or dependen* or disorder# or habituat* or misus* or (mis N0 us*) or (non N0 medical*) or nonmedical* or (non N0 prescrib*) or nonprescrib* or (non N0 prescription*) or nonprescription* or withdrawal* or (("use" or used or uses or using) N2 (illicit* or illegal*))) ) | Search modes - Boolean/Phrase | Interface - EBSCOhost Research Databases  Search Screen - Advanced Search  Database - CINAHL Plus with Full Text | 1,662 |
| S66 | TI ( (drug or drugs or substance#) N3 (abus* or addict* or dependen* or disorder# or habituat* or misus* or (mis N0 us*) or (non N0 medical*) or nonmedical* or (non N0 prescrib*) or nonprescrib* or (non N0 prescription*) or nonprescription* or withdrawal* or (("use" or used or uses or using) N2 (illicit* or illegal*))) ) OR AB ( (drug or drugs or substance#) N3 (abus* or addict* or dependen* or disorder# or habituat* or misus* or (mis N0 us*) or (non N0 medical*) or nonmedical* or (non N0 prescrib*) or nonprescrib* or (non N0 prescription*) or nonprescription* or withdrawal* or (("use" or used or uses or using) N2 (illicit* or illegal*))) ) | Search modes - Boolean/Phrase | Interface - EBSCOhost Research Databases  Search Screen - Advanced Search  Database - CINAHL Plus with Full Text | 45,397 |
| S65 | TI ( (narcotic* or opioid* or opiate*) N3 (abus* or addict* or dependen* or disorder# or habituat* or misus* or (mis N0 us*) or (non N0 medical*) or nonmedical* or (non N0 prescrib*) or nonprescrib* or (non N0 prescription*) or nonprescription* or withdrawal* or (("use" or used or uses or using) N2 (illicit* or illegal*))) ) OR AB ( (narcotic* or opioid* or opiate*) N3 (abus* or addict* or dependen* or disorder# or habituat* or misus* or (mis N0 us*) or (non N0 medical*) or nonmedical* or (non N0 prescrib*) or nonprescrib* or (non N0 prescription*) or nonprescription* or withdrawal* or (("use" or used or uses or using) N2 (illicit* or illegal*))) ) | Search modes - Boolean/Phrase | Interface - EBSCOhost Research Databases  Search Screen - Advanced Search  Database - CINAHL Plus with Full Text | 10,195 |
| S64 | TI ( alcoholis* or alcoholic* ) OR AB ( alcoholis* or alcoholic* ) OR TI ( (binge# or binging) N2 (alcohol* or drink*) ) OR AB ( (binge# or binging) N2 (alcohol* or drink*) ) OR TI ( alcohol* N3 (abus* or addict* or dependen* or disorder# or habituat* or misus* or (mis N0 us*)) ) OR AB ( alcohol* N3 (abus* or addict* or dependen* or disorder# or habituat* or misus* or (mis N0 us*)) ) | Search modes - Boolean/Phrase | Interface - EBSCOhost Research Databases  Search Screen - Advanced Search  Database - CINAHL Plus with Full Text | 34,491 |
| S63 | (MH "Substance Use Disorders+") | Search modes - Boolean/Phrase | Interface - EBSCOhost Research Databases  Search Screen - Advanced Search  Database - CINAHL Plus with Full Text | 166,205 |
| S62 | S10 OR S11 OR S12 OR S13 OR S14 OR S15 OR S16 OR S17 OR S18 OR S19 OR S20 OR S21 OR S22 OR S23 OR S24 OR S25 OR S26 OR S27 OR S28 OR S29 OR S30 OR S31 OR S32 OR S33 OR S34 OR S35 OR S36 OR S37 OR S38 OR S39 OR S40 OR S41 OR S42 OR S43 OR S44 OR S45 OR S46 OR S47 OR S48 OR S49 OR S50 OR S51 OR S52 OR S53 OR S54 OR S55 OR S56 OR S57 OR S58 OR S59 OR S60 OR S61 | Search modes - Boolean/Phrase | Interface - EBSCOhost Research Databases  Search Screen - Advanced Search  Database - CINAHL Plus with Full Text | 442,089 |
| S61 | TI ( neonat* N2 ("critical care" or "intensive care" or ICU or ICUs) N3 (discharg* or graduat* or surviv*) ) OR AB ( neonat* N2 ("critical care" or "intensive care" or ICU or ICUs) N3 (discharg* or graduat* or surviv*) ) OR TI ( post* N2 (NICU or NICUs) ) OR AB ( post* N2 (NICU or NICUs) ) OR TI ( (NICU or NICUs) N3 (discharg* or graduat* or surviv*) ) OR AB ( (NICU or NICUs) N3 (discharg* or graduat* or surviv*) ) | Search modes - Boolean/Phrase | Interface - EBSCOhost Research Databases  Search Screen - Advanced Search  Database - CINAHL Plus with Full Text | 869 |
| S60 | MH "Intensive Care Units, Neonatal" AND MH ( "Survivors" OR "Survivorship" OR "Patient Discharge" ) | Search modes - Boolean/Phrase | Interface - EBSCOhost Research Databases  Search Screen - Advanced Search  Database - CINAHL Plus with Full Text | 443 |
| S59 | TI ( (cardiac* or cardio* or heart#) N2 (abnormalit* or anomal* or atypical* or a-typical* or defect* or deficien* or deform* or impair* or malform*) ) OR AB ( (cardiac* or cardio* or heart#) N2 (abnormalit* or anomal* or atypical* or a-typical* or defect* or deficien* or deform* or impair* or malform*) ) OR TI tetralog* N2 fallot* OR AB tetralog* N2 fallot* OR TI ( (cardiac* or cardio* or heart#) N5 (congenital* or inborn* or hereditar* or inherit*) ) OR AB ( (cardiac* or cardio* or heart#) N5 (congenital* or inborn* or hereditar* or inherit*) ) | Search modes - Boolean/Phrase | Interface - EBSCOhost Research Databases  Search Screen - Advanced Search  Database - CINAHL Plus with Full Text | 19,534 |
| S58 | (MH "Heart Defects, Congenital+") | Search modes - Boolean/Phrase | Interface - EBSCOhost Research Databases  Search Screen - Advanced Search  Database - CINAHL Plus with Full Text | 27,295 |
| S57 | TI "small for gestational" N0 age# OR AB "small for gestational" N0 age# OR TI ( SGA or LBW or VLBW ) OR AB ( SGA or LBW or VLBW ) OR TI low* N0 birth N0 weight# OR AB low* N0 birth N0 weight# | Search modes - Boolean/Phrase | Interface - EBSCOhost Research Databases  Search Screen - Advanced Search  Database - CINAHL Plus with Full Text | 15,326 |
| S56 | (MH "Infant, Low Birth Weight+") | Search modes - Boolean/Phrase | Interface - EBSCOhost Research Databases  Search Screen - Advanced Search  Database - CINAHL Plus with Full Text | 14,444 |
| S55 | TI ( (preterm or "pre-term" or prematur* or (pre N0 matur*)) N3 (baby or babies or infant* or neonat* or newborn*) ) OR AB ( (preterm or "pre-term" or prematur* or (pre N0 matur*)) N3 (baby or babies or infant* or neonat* or newborn*) ) | Search modes - Boolean/Phrase | Interface - EBSCOhost Research Databases  Search Screen - Advanced Search  Database - CINAHL Plus with Full Text | 23,017 |
| S54 | (MH "Infant, Premature") | Search modes - Boolean/Phrase | Interface - EBSCOhost Research Databases  Search Screen - Advanced Search  Database - CINAHL Plus with Full Text | 23,339 |
| S53 | (MH "Hypoxia-Ischemia, Brain, Neonatal") | Search modes - Boolean/Phrase | Interface - EBSCOhost Research Databases  Search Screen - Advanced Search  Database - CINAHL Plus with Full Text | 240 |
| S52 | TI ( (neonat* or newborn*) N3 (anoxi* or asphyxi* or hypoxi* or (respiratory N0 failure*)) ) OR AB ( (neonat* or newborn*) N3 (anoxi* or asphyxi* or hypoxi* or (respiratory N0 failure*)) ) | Search modes - Boolean/Phrase | Interface - EBSCOhost Research Databases  Search Screen - Advanced Search  Database - CINAHL Plus with Full Text | 1,807 |
| S51 | (MH "Asphyxia Neonatorum") | Search modes - Boolean/Phrase | Interface - EBSCOhost Research Databases  Search Screen - Advanced Search  Database - CINAHL Plus with Full Text | 1,174 |
| S50 | TI ( "cerebral vascular" N2 (accident* or arrest* or failure* or injur* or insufficienc* or insult*) ) OR AB ( "cerebral vascular" N2 (accident* or arrest* or failure* or injur* or insufficienc* or insult*) ) OR TI ( (isch#emic N0 seizure*) or (isch#emic N0 cerebral N0 attack*) ) OR AB ( (isch#emic N0 seizure*) or (isch#emic N0 cerebral N0 attack*) ) OR TI ( apoplex* or apoplectic* ) OR AB ( apoplex* or apoplectic* ) OR TI ( (brain or cerebral) N2 insult* ) OR AB ( (brain or cerebral) N2 insult* ) | Search modes - Boolean/Phrase | Interface - EBSCOhost Research Databases  Search Screen - Advanced Search  Database - CINAHL Plus with Full Text | 1,119 |
| S49 | TI stroke# OR AB stroke# OR TI ( (cerebrovascular or "cerebro-vascular") N2 (accident* or arrest* or failure* or injur* or insufficienc* or insult*) ) OR AB ( (cerebrovascular or "cerebro-vascular") N2 (accident* or arrest* or failure* or injur* or insufficienc* or insult*) ) | Search modes - Boolean/Phrase | Interface - EBSCOhost Research Databases  Search Screen - Advanced Search  Database - CINAHL Plus with Full Text | 96,911 |
| S48 | (MH "Stroke+") | Search modes - Boolean/Phrase | Interface - EBSCOhost Research Databases  Search Screen - Advanced Search  Database - CINAHL Plus with Full Text | 70,982 |
| S47 | TI ( brain# N3 (injur* or commotio* or concuss* or damag* or trauma*) ) OR AB ( brain# N3 (injur* or commotio* or concuss* or damag* or trauma*) ) OR TI concussion* OR AB concussion* OR TI commotio OR AB commotio | Search modes - Boolean/Phrase | Interface - EBSCOhost Research Databases  Search Screen - Advanced Search  Database - CINAHL Plus with Full Text | 34,826 |
| S46 | (MH "Brain Injuries+") | Search modes - Boolean/Phrase | Interface - EBSCOhost Research Databases  Search Screen - Advanced Search  Database - CINAHL Plus with Full Text | 29,741 |
| S45 | TI ( angelman* or "happy puppet" or (puppet N0 child*) ) OR AB ( angelman* or "happy puppet" or (puppet N0 child*) ) | Search modes - Boolean/Phrase | Interface - EBSCOhost Research Databases  Search Screen - Advanced Search  Database - CINAHL Plus with Full Text | 270 |
| S44 | (MH "Angelman Syndrome") | Search modes - Boolean/Phrase | Interface - EBSCOhost Research Databases  Search Screen - Advanced Search  Database - CINAHL Plus with Full Text | 227 |
| S43 | TI "Fragile X" OR AB "Fragile X" OR TI ( FRAXA or FRAXE or "Fra(X)" ) OR AB ( FRAXA or FRAXE or "Fra(X)" ) OR TI ( "Mar (X)" or "Marker X" or "Martin-Bell" ) OR AB ( "Mar (X)" or "Marker X" or "Martin-Bell" ) OR TI "X-Linked" N2 "mental retardation" OR AB "X-Linked" N2 "mental retardation" | Search modes - Boolean/Phrase | Interface - EBSCOhost Research Databases  Search Screen - Advanced Search  Database - CINAHL Plus with Full Text | 1,123 |
| S42 | (MH "Fragile X Syndrome") | Search modes - Boolean/Phrase | Interface - EBSCOhost Research Databases  Search Screen - Advanced Search  Database - CINAHL Plus with Full Text | 997 |
| S41 | TI ( (neonat* or newborn*) N2 (abstinen* or addiction* or withdrawal*) ) OR AB ( (neonat* or newborn*) N2 (abstinen* or addiction* or withdrawal*) ) | Search modes - Boolean/Phrase | Interface - EBSCOhost Research Databases  Search Screen - Advanced Search  Database - CINAHL Plus with Full Text | 954 |
| S40 | (MH "Neonatal Abstinence Syndrome") | Search modes - Boolean/Phrase | Interface - EBSCOhost Research Databases  Search Screen - Advanced Search  Database - CINAHL Plus with Full Text | 1,115 |
| S39 | TI ( "alcohol-related" N3 (neurodevelopment* or neuro-development*) N2 (deficit* or delay* or deviation* or disabil* or disabled or disorder* or disturb* or handicap* or impair*) ) OR AB ( "alcohol-related" N3 (neurodevelopment* or neuro-development*) N2 (deficit* or delay* or deviation* or disabil* or disabled or disorder* or disturb* or handicap* or impair*) ) | Search modes - Boolean/Phrase | Interface - EBSCOhost Research Databases  Search Screen - Advanced Search  Database - CINAHL Plus with Full Text | 42 |
| S38 | TI ( (f#etal or f#etus*) N2 (alcohol N0 syndrome*) ) OR AB ( (f#etal or f#etus*) N2 (alcohol N0 syndrome*) ) OR TI FASD OR AB FASD OR TI ( "alcohol-related" N3 ((birth N0 defect*) or (birth N0 disorder*)) ) OR AB ( "alcohol-related" N3 ((birth N0 defect*) or (birth N0 disorder*)) ) | Search modes - Boolean/Phrase | Interface - EBSCOhost Research Databases  Search Screen - Advanced Search  Database - CINAHL Plus with Full Text | 1,072 |
| S37 | (MH "Fetal Alcohol Syndrome") | Search modes - Boolean/Phrase | Interface - EBSCOhost Research Databases  Search Screen - Advanced Search  Database - CINAHL Plus with Full Text | 1,851 |
| S36 | TI Down* N2 syndrome* OR AB Down* N2 syndrome* OR TI ( mongolism* or mongoloid* ) OR AB ( mongolism* or mongoloid* ) OR TI ( "trisomy 21" or "trisomy G1" or "trisomy (G)1" or "trisomy G-1" or "trisomy GM" or "trisomy G" or "21 trisomy" or "G1 trisomy" or "G(1) trisomy" or "G-1 trisomy" or "GM trisomy" or "G trisomy" ) OR AB ( "trisomy 21" or "trisomy G1" or "trisomy (G)1" or "trisomy G-1" or "trisomy GM" or "trisomy G" or "21 trisomy" or "G1 trisomy" or "G(1) trisomy" or "G-1 trisomy" or "GM trisomy" or "G trisomy" ) OR TI translocat* N1 DS OR AB translocat* N1 DS | Search modes - Boolean/Phrase | Interface - EBSCOhost Research Databases  Search Screen - Advanced Search  Database - CINAHL Plus with Full Text | 7,373 |
| S35 | (MH "Down Syndrome") | Search modes - Boolean/Phrase | Interface - EBSCOhost Research Databases  Search Screen - Advanced Search  Database - CINAHL Plus with Full Text | 7,432 |
| S34 | TI ( cognitive* N2 (atypical* or a-typical* or deficit* or delay* or deviation* or disabil* or disabled or disorder* or disturb* or dysfunction* or handicap* or impair* or retard*) ) OR AB ( cognitive* N2 (atypical* or a-typical* or deficit* or delay* or deviation* or disabil* or disabled or disorder* or disturb* or dysfunction* or handicap* or impair* or retard*) ) OR TI ( cognition N2 (atypical* or a-typical* or deficit* or delay* or deviation* or disabil* or disabled or disorder* or disturb* or dysfunction* or handicap* or impair* or retard*) ) OR AB ( cognition N2 (atypical* or a-typical* or deficit* or delay* or deviation* or disabil* or disabled or disorder* or disturb* or dysfunction* or handicap* or impair* or retard*) ) OR TI ( mental* N2 (atypical* or a-typical* or deficit* or delay* or deviation* or disabil* or disabled or disorder* or disturb* or dysfunction* or handicap* or impair* or retard*) ) OR AB ( mental* N2 (atypical* or a-typical* or deficit* or delay* or deviation* or disabil* or disabled or disorder* or disturb* or dysfunction* or handicap* or impair* or retard*) ) | Search modes - Boolean/Phrase | Interface - EBSCOhost Research Databases  Search Screen - Advanced Search  Database - CINAHL Plus with Full Text | 64,967 |
| S33 | TI ( intellectual* N2 (atypical* or a-typical* or deficit* or delay* or deviation* or disabil* or disabled or disorder* or disturb* or dysfunction* or handicap* or impair* or retard*) ) OR AB ( intellectual* N2 (atypical* or a-typical* or deficit* or delay* or deviation* or disabil* or disabled or disorder* or disturb* or dysfunction* or handicap* or impair* or retard*) ) OR TI ( brain# N2 (atypical* or a-typical* or deficit* or delay* or deviation* or disabil* or disabled or disorder* or disturb* or dysfunction* or handicap* or impair* or retard*) ) OR AB ( brain# N2 (atypical* or a-typical* or deficit* or delay* or deviation* or disabil* or disabled or disorder* or disturb* or dysfunction* or handicap* or impair* or retard*) ) | Search modes - Boolean/Phrase | Interface - EBSCOhost Research Databases  Search Screen - Advanced Search  Database - CINAHL Plus with Full Text | 18,411 |
| S32 | (MH "Intellectual Disability") | Search modes - Boolean/Phrase | Interface - EBSCOhost Research Databases  Search Screen - Advanced Search  Database - CINAHL Plus with Full Text | 21,295 |
| S31 | TI spina# N0 bifida* OR AB spina# N0 bifida* OR ( (cleft N0 spine#) or (open N0 spine#) or rachischis* or schistorrhach* or spina? dysraphia* or status dysraphicus ) | Search modes - Boolean/Phrase | Interface - EBSCOhost Research Databases  Search Screen - Advanced Search  Database - CINAHL Plus with Full Text | 1,904 |
| S30 | TI ( acrania# or craniorachischis* or diastematomyelia* or exencephal* or iniencephal* or (neurenteric N0 cyst#) or (neuroenteric N0 cyst#) or (spinal N0 dysraphism*) or ("spinal cord" N0 myelodysplasia*) or (tethered N2 cord N0 syndrome*) ) OR AB ( acrania# or craniorachischis* or diastematomyelia* or exencephal* or iniencephal* or (neurenteric N0 cyst#) or (neuroenteric N0 cyst#) or (spinal N0 dysraphism*) or ("spinal cord" N0 myelodysplasia*) or (tethered N2 cord N0 syndrome*) ) | Search modes - Boolean/Phrase | Interface - EBSCOhost Research Databases  Search Screen - Advanced Search  Database - CINAHL Plus with Full Text | 492 |
| S29 | TI neural N0 tube# N2 defect* OR AB neural N0 tube# N2 defect* | Search modes - Boolean/Phrase | Interface - EBSCOhost Research Databases  Search Screen - Advanced Search  Database - CINAHL Plus with Full Text | 1,562 |
| S28 | (MH "Neural Tube Defects+") | Search modes - Boolean/Phrase | Interface - EBSCOhost Research Databases  Search Screen - Advanced Search  Database - CINAHL Plus with Full Text | 5,826 |
| S27 | (MH "Malformations of Cortical Development+") | Search modes - Boolean/Phrase | Interface - EBSCOhost Research Databases  Search Screen - Advanced Search  Database - CINAHL Plus with Full Text | 303 |
| S26 | TI ( (brain or central* or cerebral*) N1 (paralys* or pares?s) ) OR AB ( (brain or central* or cerebral*) N1 (paralys* or pares?s) ) OR TI "encephalopathia infantilis" OR AB "encephalopathia infantilis" | Search modes - Boolean/Phrase | Interface - EBSCOhost Research Databases  Search Screen - Advanced Search  Database - CINAHL Plus with Full Text | 91 |
| S25 | TI ( (brain# or central*) N1 palsy ) OR AB ( (brain# or central*) N1 palsy ) | Search modes - Boolean/Phrase | Interface - EBSCOhost Research Databases  Search Screen - Advanced Search  Database - CINAHL Plus with Full Text | 80 |
| S24 | TI ( (cerebral N1 palsy) or (diplegia* N1 spastic) or (little N1 disease) ) OR AB ( (cerebral N1 palsy) or (diplegia* N1 spastic) or (little N1 disease) ) | Search modes - Boolean/Phrase | Interface - EBSCOhost Research Databases  Search Screen - Advanced Search  Database - CINAHL Plus with Full Text | 13,503 |
| S23 | (MH "Cerebral Palsy") | Search modes - Boolean/Phrase | Interface - EBSCOhost Research Databases  Search Screen - Advanced Search  Database - CINAHL Plus with Full Text | 12,322 |
| S22 | TI ( (language* or auditor* or semantic* or speech or speak* or talk* or verbal*) N3 (atypical* or a-typical* or deficit* or delay* or disabil* or disabled or disorder* or disturb* or dysfunction* or handicap* or impair* or retard*) ) OR AB ( (language* or auditor* or semantic* or speech or speak* or talk* or verbal*) N3 (atypical* or a-typical* or deficit* or delay* or disabil* or disabled or disorder* or disturb* or dysfunction* or handicap* or impair* or retard*) ) | Search modes - Boolean/Phrase | Interface - EBSCOhost Research Databases  Search Screen - Advanced Search  Database - CINAHL Plus with Full Text | 15,754 |
| S21 | (MH "Language Disorders+") | Search modes - Boolean/Phrase | Interface - EBSCOhost Research Databases  Search Screen - Advanced Search  Database - CINAHL Plus with Full Text | 22,214 |
| S20 | TI ( alexia* or alexic* or dyslexia* or dyslexic* or (reading N0 disab*) or "word blindness" or (verbal* N1 agnosi*) ) OR AB ( alexia* or alexic* or dyslexia* or dyslexic* or (reading N0 disab*) or "word blindness" or (verbal* N1 agnosi*) ) | Search modes - Boolean/Phrase | Interface - EBSCOhost Research Databases  Search Screen - Advanced Search  Database - CINAHL Plus with Full Text | 2,659 |
| S19 | TI ( learning N3 (atypical* or a-typical* or deficit* or delay* or disabil* or disabled or disorder* or disturb* or dysfunction* or handicap* or impair* or retard*) ) OR AB ( learning N3 (atypical* or a-typical* or deficit* or delay* or disabil* or disabled or disorder* or disturb* or dysfunction* or handicap* or impair* or retard*) ) OR TI ( scholastic* N3 (atypical* or a-typical* or deficit* or delay* or disabil* or disabled or disorder* or disturb* or dysfunction* or handicap* or impair* or retard*) ) OR AB ( scholastic* N3 (atypical* or a-typical* or deficit* or delay* or disabil* or disabled or disorder* or disturb* or dysfunction* or handicap* or impair* or retard*) ) | Search modes - Boolean/Phrase | Interface - EBSCOhost Research Databases  Search Screen - Advanced Search  Database - CINAHL Plus with Full Text | 13,160 |
| S18 | (MH "Learning Disorders+") | Search modes - Boolean/Phrase | Interface - EBSCOhost Research Databases  Search Screen - Advanced Search  Database - CINAHL Plus with Full Text | 10,154 |
| S17 | TI ADHD OR AB ADHD OR TI ( (hyperkinetic or kanner*) N0 syndrome* ) OR AB ( (hyperkinetic or kanner*) N0 syndrome* ) | Search modes - Boolean/Phrase | Interface - EBSCOhost Research Databases  Search Screen - Advanced Search  Database - CINAHL Plus with Full Text | 12,926 |
| S16 | TI attention N0 deficit* OR AB attention N0 deficit* | Search modes - Boolean/Phrase | Interface - EBSCOhost Research Databases  Search Screen - Advanced Search  Database - CINAHL Plus with Full Text | 7,986 |
| S15 | (MH "Attention Deficit Hyperactivity Disorder") | Search modes - Boolean/Phrase | Interface - EBSCOhost Research Databases  Search Screen - Advanced Search  Database - CINAHL Plus with Full Text | 16,665 |
| S14 | TI ( autis* or asperger* ) OR AB ( autis* or asperger* ) | Search modes - Boolean/Phrase | Interface - EBSCOhost Research Databases  Search Screen - Advanced Search  Database - CINAHL Plus with Full Text | 26,725 |
| S13 | (MH "Child Development Disorders, Pervasive+") | Search modes - Boolean/Phrase | Interface - EBSCOhost Research Databases  Search Screen - Advanced Search  Database - CINAHL Plus with Full Text | 26,896 |
| S12 | TI global N2 delay* OR AB global N2 delay* | Search modes - Boolean/Phrase | Interface - EBSCOhost Research Databases  Search Screen - Advanced Search  Database - CINAHL Plus with Full Text | 377 |
| S11 | TI ( development* N2 (abnormal* or atypical* or a-typical* or deficit* or delay* or deviation* or disabil* or disabled or disorder* or disturb* or handicap* or impair*) ) OR AB ( development* N2 (abnormal* or atypical* or a-typical* or deficit* or delay* or deviation* or disabil* or disabled or disorder* or disturb* or handicap* or impair*) ) | Search modes - Boolean/Phrase | Interface - EBSCOhost Research Databases  Search Screen - Advanced Search  Database - CINAHL Plus with Full Text | 22,143 |
| S10 | (MH "Developmental Disabilities") | Search modes - Boolean/Phrase | Interface - EBSCOhost Research Databases  Search Screen - Advanced Search  Database - CINAHL Plus with Full Text | 10,119 |
| S9 | S1 OR S2 OR S3 OR S4 OR S5 OR S6 OR S7 OR S8 | Search modes - Boolean/Phrase | Interface - EBSCOhost Research Databases  Search Screen - Advanced Search  Database - CINAHL Plus with Full Text | 28,158 |
| S8 | TI coronavirus* or (corona N0 virus*) | Search modes - Boolean/Phrase | Interface - EBSCOhost Research Databases  Search Screen - Advanced Search  Database - CINAHL Plus with Full Text | 3,381 |
| S7 | TX ( (novel N0 coronavirus*) or ("novel corona" N0 virus*) ) OR TX ( (coronavirus* or (corona" N0 virus*)) N2 "2019" ) OR TX ( (coronavirus* or (corona" N0 virus*)) N2 "19" ) OR TX ( "coronavirus 2" or "corona virus 2" ) | Search modes - Boolean/Phrase | Interface - EBSCOhost Research Databases  Search Screen - Advanced Search  Database - CINAHL Plus with Full Text | 15,523 |
| S6 | TX "2019-nCoV" OR TX ( nCoV or "n-CoV" ) OR TX "HCoV-19" OR TX ( "SARS-CoV-2" or "SARS-CoV2" or "SARSCoV-2" or SARSCoV2 ) | Search modes - Boolean/Phrase | Interface - EBSCOhost Research Databases  Search Screen - Advanced Search  Database - CINAHL Plus with Full Text | 3,481 |
| S5 | TX Wuhan N0 virus* | Search modes - Boolean/Phrase | Interface - EBSCOhost Research Databases  Search Screen - Advanced Search  Database - CINAHL Plus with Full Text | 10 |
| S4 | TX ( coronavirus* and (hubei or wuhan or beijing or shanghai) ) OR TX ( (corona N0 virus*) and (hubei or wuhan or beijing or shanghai) ) | Search modes - Boolean/Phrase | Interface - EBSCOhost Research Databases  Search Screen - Advanced Search  Database - CINAHL Plus with Full Text | 1,665 |
| S3 | TX "COVID-19" or COVID19 | Search modes - Boolean/Phrase | Interface - EBSCOhost Research Databases  Search Screen - Advanced Search  Database - CINAHL Plus with Full Text | 23,657 |
| S2 | (MH "Coronavirus Infections") | Search modes - Boolean/Phrase | Interface - EBSCOhost Research Databases  Search Screen - Advanced Search  Database - CINAHL Plus with Full Text | 8,433 |
| S1 | (MH "Coronavirus") | Search modes - Boolean/Phrase | Interface - EBSCOhost Research Databases  Search Screen - Advanced Search  Database - CINAHL Plus with Full Text | 866 |

Web of Science

| # 30 | [201](https://apps-webofknowledge-com.myaccess.library.utoronto.ca/summary.do?product=WOS&doc=1&qid=31&SID=6FDTHsJqRwm6GiRNVeY&search_mode=CombineSearches&update_back2search_link_param=yes) | #28  OR  #22  **Refined by:** **PUBLICATION YEARS:** ( 2020 )  Indexes=SCI-EXPANDED, SSCI, A&HCI, CPCI-S, CPCI-SSH, BKCI-S, BKCI-SSH, ESCI Timespan=1900-2020 |
| --- | --- | --- |
| # 29 | [202](https://apps-webofknowledge-com.myaccess.library.utoronto.ca/summary.do?product=WOS&doc=1&qid=30&SID=6FDTHsJqRwm6GiRNVeY&search_mode=CombineSearches&update_back2search_link_param=yes) | #28  OR  #22  Indexes=SCI-EXPANDED, SSCI, A&HCI, CPCI-S, CPCI-SSH, BKCI-S, BKCI-SSH, ESCI Timespan=1900-2020 |
| # 28 | 0 | #27  AND  #4  Indexes=SCI-EXPANDED, SSCI, A&HCI, CPCI-S, CPCI-SSH, BKCI-S, BKCI-SSH, ESCI Timespan=1900-2020 |
| # 27 | [5,116](https://apps-webofknowledge-com.myaccess.library.utoronto.ca/summary.do?product=WOS&doc=1&qid=28&SID=6FDTHsJqRwm6GiRNVeY&search_mode=CombineSearches&update_back2search_link_param=yes) | #26  AND  #25  Indexes=SCI-EXPANDED, SSCI, A&HCI, CPCI-S, CPCI-SSH, BKCI-S, BKCI-SSH, ESCI Timespan=1900-2020 |
| # 26 | [805,394](https://apps-webofknowledge-com.myaccess.library.utoronto.ca/summary.do?product=WOS&doc=1&qid=27&SID=6FDTHsJqRwm6GiRNVeY&search_mode=GeneralSearch&update_back2search_link_param=yes) | **TOPIC:**  (before NEAR/3 (birth* or childbirth* or "child birth") )  *OR*  **TOPIC:**  (prior NEAR/3 (birth* or childbirth* or "child birth") )  *OR*  **TOPIC:**  (baby or babies or infant or infants or infanc* or neonat* or newborn*)  Indexes=SCI-EXPANDED, SSCI, A&HCI, CPCI-S, CPCI-SSH, BKCI-S, BKCI-SSH, ESCI Timespan=1900-2020 |
| # 25 | [299,959](https://apps-webofknowledge-com.myaccess.library.utoronto.ca/summary.do?product=WOS&doc=1&qid=26&SID=6FDTHsJqRwm6GiRNVeY&search_mode=CombineSearches&update_back2search_link_param=yes) | #24  OR  #23  Indexes=SCI-EXPANDED, SSCI, A&HCI, CPCI-S, CPCI-SSH, BKCI-S, BKCI-SSH, ESCI Timespan=1900-2020 |
| # 24 | [144,554](https://apps-webofknowledge-com.myaccess.library.utoronto.ca/summary.do?product=WOS&doc=1&qid=25&SID=6FDTHsJqRwm6GiRNVeY&search_mode=GeneralSearch&update_back2search_link_param=yes) | **TOPIC:**  (drug NEAR/3 (abus* or addict* or dependen* or disorder* or habituat* or misus* or mis-us*) )  *OR*  **TOPIC:**  (drugs NEAR/3 (abus* or addict* or dependen* or disorder* or habituat* or misus* or mis-us*) )  *OR*  **TOPIC:**  (substance* NEAR/3 (abus* or addict* or dependen* or disorder* or habituat* or misus* or mis-us*) )  *OR*  **TOPIC:**  (heroin* NEAR/3 (abus* or addict* or dependen* or disorder* or habituat* or misus* or mis-us*) )  *OR*  **TOPIC:**  (hydrocodone NEAR/3 (abus* or addict* or dependen* or disorder* or habituat* or misus* or mis-us* or non-medical* or nonmedical* or non-prescrib* or nonprescrib* or non-prescription* or nonprescription* or withdrawal*) )  *OR*  **TOPIC:**  (fentanyl NEAR/3 (abus* or addict* or dependen* or disorder* or habituat* or misus* or mis-us* or non-medical* or nonmedical* or non-prescrib* or nonprescrib* or non-prescription* or nonprescription* or withdrawal*) )  *OR*  **TOPIC:**  (morphine NEAR/3 (abus* or addict* or dependen* or disorder* or habituat* or misus* or mis-us* or non-medical* or nonmedical* or non-prescrib* or nonprescrib* or non-prescription* or nonprescription* or withdrawal*) )  *OR*  **TOPIC:**  (oxycodone NEAR/3 (abus* or addict* or dependen* or disorder* or habituat* or misus* or mis-us* or non-medical* or nonmedical* or non-prescrib* or nonprescrib* or non-prescription* or nonprescription* or withdrawal*) )  *OR*  **TOPIC:**  (oxycontin NEAR/3 (abus* or addict* or dependen* or disorder* or habituat* or misus* or mis-us* or non-medical* or nonmedical* or non-prescrib* or nonprescrib* or non-prescription* or nonprescription* or withdrawal*) )  Indexes=SCI-EXPANDED, SSCI, A&HCI, CPCI-S, CPCI-SSH, BKCI-S, BKCI-SSH, ESCI Timespan=1900-2020 |
| # 23 | [187,244](https://apps-webofknowledge-com.myaccess.library.utoronto.ca/summary.do?product=WOS&doc=1&qid=24&SID=6FDTHsJqRwm6GiRNVeY&search_mode=GeneralSearch&update_back2search_link_param=yes) | **TOPIC:**  (alcoholis* or alcoholic*)  *OR*  **TOPIC:**  (binge* NEAR/2 (alcohol* or drink*) )  *OR*  **TOPIC:**  (binging NEAR/2 (alcohol* or drink*) )  *OR*  **TOPIC:**  (alcohol* NEAR/3 (abus* or addict* or dependen* or disorder* or habituat* or misus* or mis-us*) )  *OR*  **TOPIC:**  (alcohol* NEAR/3 (abus* or addict* or dependen* or disorder* or habituat* or misus* or mis-us*) )  *OR*  **TOPIC:**  (narcotic* NEAR/3 (abus* or addict* or dependen* or disorder* or habituat* or misus* or mis-us*) )  *OR*  **TOPIC:**  (opioid* NEAR/3 (abus* or addict* or dependen* or disorder* or habituat* or misus* or mis-us*) )  *OR*  **TOPIC:**  (opiate* NEAR/3 (abus* or addict* or dependen* or disorder* or habituat* or misus* or mis-us*) )  Indexes=SCI-EXPANDED, SSCI, A&HCI, CPCI-S, CPCI-SSH, BKCI-S, BKCI-SSH, ESCI Timespan=1900-2020 |
| # 22 | [202](https://apps-webofknowledge-com.myaccess.library.utoronto.ca/summary.do?product=WOS&doc=1&qid=23&SID=6FDTHsJqRwm6GiRNVeY&search_mode=CombineSearches&update_back2search_link_param=yes) | #20  AND  #19  **Refined by:** **PUBLICATION YEARS:** ( 2020 OR 2019 )  Indexes=SCI-EXPANDED, SSCI, A&HCI, CPCI-S, CPCI-SSH, BKCI-S, BKCI-SSH, ESCI Timespan=1900-2020 |
| # 21 | [212](https://apps-webofknowledge-com.myaccess.library.utoronto.ca/summary.do?product=WOS&doc=1&qid=22&SID=6FDTHsJqRwm6GiRNVeY&search_mode=CombineSearches&update_back2search_link_param=yes) | #20  AND  #19  Indexes=SCI-EXPANDED, SSCI, A&HCI, CPCI-S, CPCI-SSH, BKCI-S, BKCI-SSH, ESCI Timespan=1900-2020 |
| # 20 | [2,924,348](https://apps-webofknowledge-com.myaccess.library.utoronto.ca/summary.do?product=WOS&doc=1&qid=21&SID=6FDTHsJqRwm6GiRNVeY&search_mode=GeneralSearch&update_back2search_link_param=yes) | **TOPIC:**  (baby or babies or infant or infants or infanc* or neonat* or newborn* or preschool* or pre-school* or toddler*)  *OR*  **TOPIC:**  (adolescen* or child* or pre-adolescen* or preteen* or pre-teen* or school-age* or teen or teens or teenager* or youth*)  Indexes=SCI-EXPANDED, SSCI, A&HCI, CPCI-S, CPCI-SSH, BKCI-S, BKCI-SSH, ESCI Timespan=1900-2020 |
| # 19 | [1,163](https://apps-webofknowledge-com.myaccess.library.utoronto.ca/summary.do?product=WOS&doc=1&qid=20&SID=6FDTHsJqRwm6GiRNVeY&search_mode=CombineSearches&update_back2search_link_param=yes) | #18  AND  #4  Indexes=SCI-EXPANDED, SSCI, A&HCI, CPCI-S, CPCI-SSH, BKCI-S, BKCI-SSH, ESCI Timespan=1900-2020 |
| # 18 | [1,317,857](https://apps-webofknowledge-com.myaccess.library.utoronto.ca/summary.do?product=WOS&doc=1&qid=19&SID=6FDTHsJqRwm6GiRNVeY&search_mode=CombineSearches&update_back2search_link_param=yes) | #17  OR  #16  OR  #15  OR  #14  OR  #13  OR  #12  OR  #11  OR  #10  OR  #9  OR  #8  OR  #7  OR  #6  OR  #5  Indexes=SCI-EXPANDED, SSCI, A&HCI, CPCI-S, CPCI-SSH, BKCI-S, BKCI-SSH, ESCI Timespan=1900-2020 |
| # 17 | [1,299](https://apps-webofknowledge-com.myaccess.library.utoronto.ca/summary.do?product=WOS&doc=1&qid=18&SID=6FDTHsJqRwm6GiRNVeY&search_mode=GeneralSearch&update_back2search_link_param=yes) | **TOPIC:**  ("neonatal intensive care" NEAR/3 (discharg* or graduat* or surviv*) )  *OR*  **TOPIC:**  ("neonatal critical care" NEAR/3 (discharg* or graduat* or surviv*) )  *OR*  **TOPIC:**  ("neonatal ICU" NEAR/3 (discharg* or graduat* or surviv*) )  *OR*  **TOPIC:**  ("neonatal ICUs" NEAR/3 (discharg* or graduat* or surviv*) )  *OR*  **TOPIC:**  (post* NEAR/2 (NICU or NICUs) )  *OR*  **TOPIC:**  (NICU NEAR/3 (discharg* or graduat* or surviv*) )  *OR*  **TOPIC:**  (NICUs NEAR/3 (discharg* or graduat* or surviv*) )  Indexes=SCI-EXPANDED, SSCI, A&HCI, CPCI-S, CPCI-SSH, BKCI-S, BKCI-SSH, ESCI Timespan=1900-2020 |
| # 16 | [101,209](https://apps-webofknowledge-com.myaccess.library.utoronto.ca/summary.do?product=WOS&doc=1&qid=17&SID=6FDTHsJqRwm6GiRNVeY&search_mode=GeneralSearch&update_back2search_link_param=yes) | **TOPIC:**  (cardiac* NEAR/2 (abnormalit* or anomal* or atypical* or a-typical* or defect* or deficien* or deform* or impair* or malform*) )  *OR*  **TOPIC:**  (cardio* NEAR/2 (abnormalit* or anomal* or atypical* or a-typical* or defect* or deficien* or deform* or impair* or malform*) )  *OR*  **TOPIC:**  (heart* NEAR/2 (abnormalit* or anomal* or atypical* or a-typical* or defect* or deficien* or deform* or impair* or malform*) )  *OR*  **TOPIC:**  (tetralog* NEAR/2 fallot*)  *OR*  **TOPIC:**  (cardiac* NEAR/5 (congenital* or inborn* or hereditar* or inherit*) )  *OR*  **TOPIC:**  (cardio* NEAR/5 (congenital* or inborn* or hereditar* or inherit*) )  *OR*  **TOPIC:**  (heart* NEAR/5 (congenital* or inborn* or hereditar* or inherit*) )  Indexes=SCI-EXPANDED, SSCI, A&HCI, CPCI-S, CPCI-SSH, BKCI-S, BKCI-SSH, ESCI Timespan=1900-2020 |
| # 15 | [22,278](https://apps-webofknowledge-com.myaccess.library.utoronto.ca/summary.do?product=WOS&doc=1&qid=16&SID=6FDTHsJqRwm6GiRNVeY&search_mode=GeneralSearch&update_back2search_link_param=yes) | **TOPIC:**  ("small for gestational age" or "small for gestational ages")  *OR*  **TOPIC:**  (SGA or LBW or VLBW)  *OR*  **TOPIC:**  (low* W/0 birth W/0 weight*)  Indexes=SCI-EXPANDED, SSCI, A&HCI, CPCI-S, CPCI-SSH, BKCI-S, BKCI-SSH, ESCI Timespan=1900-2020 |
| # 14 | [81,978](https://apps-webofknowledge-com.myaccess.library.utoronto.ca/summary.do?product=WOS&doc=1&qid=15&SID=6FDTHsJqRwm6GiRNVeY&search_mode=GeneralSearch&update_back2search_link_param=yes) | **TOPIC:**  (neonat* NEAR/3 (anoxi* or asphyxi* or hypoxi* or "respiratory failure" or "respiratory failures") )  *OR*  **TOPIC:**  (newborn* NEAR/3 (anoxi* or asphyxi* or hypoxi* or "respiratory failure" or "respiratory failures") )  *OR*  **TOPIC:**  (preterm NEAR/3 (baby or babies or infant* or neonat* or newborn*) )  *OR*  **TOPIC:**  ("pre-term" NEAR/3 (baby or babies or infant* or neonat* or newborn*) )  *OR*  **TOPIC:**  (prematur* NEAR/3 (baby or babies or infant* or neonat* or newborn*) )  *OR*  **TOPIC:**  (pre-matur* NEAR/3 (baby or babies or infant* or neonat* or newborn*) )  Indexes=SCI-EXPANDED, SSCI, A&HCI, CPCI-S, CPCI-SSH, BKCI-S, BKCI-SSH, ESCI Timespan=1900-2020 |
| # 13 | [504,173](https://apps-webofknowledge-com.myaccess.library.utoronto.ca/summary.do?product=WOS&doc=1&qid=14&SID=6FDTHsJqRwm6GiRNVeY&search_mode=GeneralSearch&update_back2search_link_param=yes) | **TOPIC:**  (brain* NEAR/3 (injur* or commotio* or concuss* or damag* or trauma*) )  *OR*  **TOPIC:**  (concussion* or commotio)  *OR*  **TOPIC:**  (stroke or strokes)  *OR*  **TOPIC:**  (cerebrovascular NEAR/2 (accident* or arrest* or failure* or injur* or insufficienc* or insult*) )  *OR*  **TOPIC:**  ("cerebro-vascular" NEAR/2 (accident* or arrest* or failure* or injur* or insufficienc* or insult*) )  *OR*  **TOPIC:**  ("cerebal vascular" NEAR/2 (accident* or arrest* or failure* or injur* or insufficienc* or insult*) )  Indexes=SCI-EXPANDED, SSCI, A&HCI, CPCI-S, CPCI-SSH, BKCI-S, BKCI-SSH, ESCI Timespan=1900-2020 |
| # 12 | [17,636](https://apps-webofknowledge-com.myaccess.library.utoronto.ca/summary.do?product=WOS&doc=1&qid=13&SID=6FDTHsJqRwm6GiRNVeY&search_mode=AdvancedSearch&update_back2search_link_param=yes) | TS=(neonat*  NEAR/2  (abstinen* or addiction* or withdrawal*) )  OR  TS=(newborn  NEAR/2  (abstinen* or addiction* or withdrawal*) )  OR  TS=("Fragile  X"  or  FRAXA  or  FRAXE  or  "Fra(X)"  or  "Mar  (X) "  or  "Marker  X"  or  "Martin-Bell")  OR  TS=("X-Linked"  NEAR/2  "mental  retardation")  OR  TS=(angelman*  or  "happy  puppet"  or  (puppet W/0 child*) )  Indexes=SCI-EXPANDED, SSCI, A&HCI, CPCI-S, CPCI-SSH, BKCI-S, BKCI-SSH, ESCI Timespan=1900-2020 |
| # 11 | [162](https://apps-webofknowledge-com.myaccess.library.utoronto.ca/summary.do?product=WOS&doc=1&qid=12&SID=6FDTHsJqRwm6GiRNVeY&search_mode=GeneralSearch&update_back2search_link_param=yes) | **TOPIC:**  ("alcohol-related")  *AND*  **TOPIC:**  (neurodevelopment* NEAR/2 (deficit* or delay* or deviation* or disabil* or disabled or disorder* or disturb* or handicap* or impair*) )  Indexes=SCI-EXPANDED, SSCI, A&HCI, CPCI-S, CPCI-SSH, BKCI-S, BKCI-SSH, ESCI Timespan=1900-2020 |
| # 10 | [39,290](https://apps-webofknowledge-com.myaccess.library.utoronto.ca/summary.do?product=WOS&doc=1&qid=11&SID=6FDTHsJqRwm6GiRNVeY&search_mode=GeneralSearch&update_back2search_link_param=yes) | **TOPIC:**  (Down* NEAR/2 syndrome*)  *OR*  **TOPIC:**  (mongolism* or mongoloid*)  *OR*  **TOPIC:**  ("trisomy 21" or "trisomy G1" or "trisomy (G) 1"  or  "trisomy  G-1"  or  "trisomy  GM"  or  "trisomy  G"  or  "21  trisomy"  or  "G1  trisomy"  or  "G(1)  trisomy"  or  "G-1  trisomy"  or  "GM  trisomy"  or  "G  trisomy")  *OR*  **TOPIC:**  ((fetal or foetal or fetus or foetus)  N2  "alcohol  syndrome")  *OR*  **TOPIC:**  ((fetal or foetal or fetus or foetus)  N2  "alcohol  syndromes")  *OR*  **TOPIC:**  (FASD)  *OR*  **TOPIC:**  ("alcohol-related" NEAR/3 ("birth defect" or "birth defects" or "birth disorder" or "birth disorders") )  Indexes=SCI-EXPANDED, SSCI, A&HCI, CPCI-S, CPCI-SSH, BKCI-S, BKCI-SSH, ESCI Timespan=1900-2020 |
| # 9 | [336,126](https://apps-webofknowledge-com.myaccess.library.utoronto.ca/summary.do?product=WOS&doc=1&qid=10&SID=6FDTHsJqRwm6GiRNVeY&search_mode=GeneralSearch&update_back2search_link_param=yes) | **TOPIC:**  (intellectual* NEAR/2 (atypical* or a-typical* or deficit* or delay* or deviation* or disabil* or disabled or disorder* or disturb* or dysfunction* or handicap* or impair* or retard*) )  *OR*  **TOPIC:**  (brain* NEAR/2 (atypical* or a-typical* or deficit* or delay* or deviation* or disabil* or disabled or disorder* or disturb* or dysfunction* or handicap* or impair* or retard*) )  *OR*  **TOPIC:**  (cognitiv* NEAR/2 (atypical* or a-typical* or deficit* or delay* or deviation* or disabil* or disabled or disorder* or disturb* or dysfunction* or handicap* or impair* or retard*) )  *OR*  **TOPIC:**  (cognition NEAR/2 (atypical* or a-typical* or deficit* or delay* or deviation* or disabil* or disabled or disorder* or disturb* or dysfunction* or handicap* or impair* or retard*) )  *OR*  **TOPIC:**  (mental* NEAR/2 (atypical* or a-typical* or deficit* or delay* or deviation* or disabil* or disabled or disorder* or disturb* or dysfunction* or handicap* or impair* or retard*) )  Indexes=SCI-EXPANDED, SSCI, A&HCI, CPCI-S, CPCI-SSH, BKCI-S, BKCI-SSH, ESCI Timespan=1900-2020 |
| # 8 | [44,975](https://apps-webofknowledge-com.myaccess.library.utoronto.ca/summary.do?product=WOS&doc=1&qid=8&SID=6FDTHsJqRwm6GiRNVeY&search_mode=GeneralSearch&update_back2search_link_param=yes) | **TOPIC:**  ((cerebral NEAR/1 palsy)  or  (diplegia* NEAR/1 spastic)  or  (little NEAR/1 disease) )  *OR*  **TOPIC:**  ((brain* or central*)  NEAR/1  palsy)  *OR*  **TOPIC:**  ((brain* or central* or cerebral*)  NEAR/1  paralys*)  *OR*  **TOPIC:**  ((brain* or central* or cerebral*)  NEAR/1  paresis)  *OR*  **TOPIC:**  ((brain* or central* or cerebral*)  NEAR/1  pareses)  *OR*  **TOPIC:**  (neural W/0 tube* NEAR/2 defect*)  *OR*  **TOPIC:**  (acrania* or craniorachischis* or diastematomyelia* or exencephal* or iniencephal* or (neurenteric W/0 cyst*)  or  (neuroenteric W/0 cyst*)  or  (spinal W/0 dysraphism*)  or  ("spinal cord" W/0 myelodysplasia*)  or  (tethered NEAR/2 cord W/0 syndrome*) )  *OR*  **TOPIC:**  ((spina* W/0 bifida*)  OR  ( cleft W/0 spine*)  or  (open W/0 spine*)  or  rachischis*  or  schistorrhach*  or  (spina* W/0 dysraphia*)  or  (status W/0 dysraphicus) )  Indexes=SCI-EXPANDED, SSCI, A&HCI, CPCI-S, CPCI-SSH, BKCI-S, BKCI-SSH, ESCI Timespan=1900-2020 |
| # 7 | [48,189](https://apps-webofknowledge-com.myaccess.library.utoronto.ca/summary.do?product=WOS&doc=1&qid=7&SID=6FDTHsJqRwm6GiRNVeY&search_mode=GeneralSearch&update_back2search_link_param=yes) | **TOPIC:**  (language NEAR/3 (atypical* or a-typical* or deficit* or delay* or disabil* or disabled or disorder* or disturb* or dysfunction* or handicap* or impair* or retard*) )  *OR*  **TOPIC:**  (auditor* NEAR/3 (atypical* or a-typical* or deficit* or delay* or disabil* or disabled or disorder* or disturb* or dysfunction* or handicap* or impair* or retard*) )  *OR*  **TOPIC:**  (semantic* NEAR/3 (atypical* or a-typical* or deficit* or delay* or disabil* or disabled or disorder* or disturb* or dysfunction* or handicap* or impair* or retard*) )  *OR*  **TOPIC:**  (speech NEAR/3 (atypical* or a-typical* or deficit* or delay* or disabil* or disabled or disorder* or disturb* or dysfunction* or handicap* or impair* or retard*) )  *OR*  **TOPIC:**  (speak* NEAR/3 (atypical* or a-typical* or deficit* or delay* or disabil* or disabled or disorder* or disturb* or dysfunction* or handicap* or impair* or retard*) )  *OR*  **TOPIC:**  (talk* NEAR/3 (atypical* or a-typical* or deficit* or delay* or disabil* or disabled or disorder* or disturb* or dysfunction* or handicap* or impair* or retard*) )  *OR*  **TOPIC:**  (verbal* NEAR/3 (atypical* or a-typical* or deficit* or delay* or disabil* or disabled or disorder* or disturb* or dysfunction* or handicap* or impair* or retard*) )  Indexes=SCI-EXPANDED, SSCI, A&HCI, CPCI-S, CPCI-SSH, BKCI-S, BKCI-SSH, ESCI Timespan=1900-2020 |
| # 6 | [61,201](https://apps-webofknowledge-com.myaccess.library.utoronto.ca/summary.do?product=WOS&doc=1&qid=6&SID=6FDTHsJqRwm6GiRNVeY&search_mode=GeneralSearch&update_back2search_link_param=yes) | **TOPIC:**  (learning NEAR/3 (atypical* or a-typical* or deficit* or delay* or disabil* or disabled or disorder* or disturb* or dysfunction* or handicap* or impair* or retard*) )  *OR*  **TOPIC:**  (scholastic* NEAR/3 (atypical* or a-typical* or deficit* or delay* or disabil* or disabled or disorder* or disturb* or dysfunction* or handicap* or impair* or retard*) )  *OR*  **TOPIC:**  (alexia* or alexic* or dyslexia* or dyslexic* or (reading W/0 disab*)  or  "word  blindness"  or  (verbal* NEAR/1 agnosi*) )  Indexes=SCI-EXPANDED, SSCI, A&HCI, CPCI-S, CPCI-SSH, BKCI-S, BKCI-SSH, ESCI Timespan=1900-2020 |
| # 5 | [222,358](https://apps-webofknowledge-com.myaccess.library.utoronto.ca/summary.do?product=WOS&doc=1&qid=5&SID=6FDTHsJqRwm6GiRNVeY&search_mode=GeneralSearch&update_back2search_link_param=yes) | **TOPIC:**  (development* NEAR/2 (abnormal* or atypical* or a-typical* or deficit* or delay* or deviation* or disabil* or disabled or disorder* or disturb* or handicap* or impair*) )  *OR*  **TOPIC:**  (global NEAR/2 delay*)  *OR*  **TOPIC:**  (autis* or asperger*)  *OR*  **TOPIC:**  (attention NEAR/0 deficit*)  *OR*  **TOPIC:**  (ADHD)  *OR*  **TOPIC:**  ((hyperkinetic or kanner*)  NEAR/0  syndrome*)  Indexes=SCI-EXPANDED, SSCI, A&HCI, CPCI-S, CPCI-SSH, BKCI-S, BKCI-SSH, ESCI Timespan=1900-2020 |
| # 4 | [57,868](https://apps-webofknowledge-com.myaccess.library.utoronto.ca/summary.do?product=WOS&doc=1&qid=4&SID=6FDTHsJqRwm6GiRNVeY&search_mode=CombineSearches&update_back2search_link_param=yes) | #3  OR  #2  OR  #1  Indexes=SCI-EXPANDED, SSCI, A&HCI, CPCI-S, CPCI-SSH, BKCI-S, BKCI-SSH, ESCI Timespan=1900-2020 |
| # 3 | [13,597](https://apps-webofknowledge-com.myaccess.library.utoronto.ca/summary.do?product=WOS&doc=1&qid=3&SID=6FDTHsJqRwm6GiRNVeY&search_mode=GeneralSearch&update_back2search_link_param=yes) | **TITLE:**  (coronavirus* or (corona W/0 virus*) )  Indexes=SCI-EXPANDED, SSCI, A&HCI, CPCI-S, CPCI-SSH, BKCI-S, BKCI-SSH, ESCI Timespan=1900-2020 |
| # 2 | [14,382](https://apps-webofknowledge-com.myaccess.library.utoronto.ca/summary.do?product=WOS&doc=1&qid=2&SID=6FDTHsJqRwm6GiRNVeY&search_mode=GeneralSearch&update_back2search_link_param=yes) | **TOPIC:**  (coronavirus* NEAR/2 "2019")  *OR*  **TOPIC:**  ((corona W/0 virus*)  NEAR/2  "2019")  *OR*  **TOPIC:**  (coronavirus* NEAR/2 "19")  *OR*  **TOPIC:**  ((corona W/0 virus*)  NEAR/2  "19")  *OR*  **TOPIC:**  ((novel W/0 coronavirus*)  or  ("novel corona" W/0 virus*) )  *OR*  **TOPIC:**  ("coronavirus 2" or "corona virus 2")  Indexes=SCI-EXPANDED, SSCI, A&HCI, CPCI-S, CPCI-SSH, BKCI-S, BKCI-SSH, ESCI Timespan=1900-2020 |
| # 1 | [49,208](https://apps-webofknowledge-com.myaccess.library.utoronto.ca/summary.do?product=WOS&doc=1&qid=1&SID=6FDTHsJqRwm6GiRNVeY&search_mode=GeneralSearch&update_back2search_link_param=yes) | **TOPIC:**  ("COVID-19" or COVID19)  *OR*  **TOPIC:**  (coronavirus* and (hubei or wuhan or beijing or shanghai) )  *OR*  **TOPIC:**  (Wuhan W/0 virus*)  *OR*  **TOPIC:**  ("2019-nCoV")  *OR*  **TOPIC:**  (nCoV or "n-CoV")  *OR*  **TOPIC:**  ("HCoV-19")  *OR*  **TOPIC:**  ("SARS-CoV-2" or "SARS-CoV2" or "SARSCoV-2" or SARSCoV2)  Indexes=SCI-EXPANDED, SSCI, A&HCI, CPCI-S, CPCI-SSH, BKCI-S, BKCI-SSH, ESCI Timespan=1900-2020 |

Cochrane Special Collection:

<https://www.cochranelibrary.com/collections/doi/SC000040/full>

*Nothing relevant to this topic 2020 Oct 31*

Cochrane

Database: EBM Reviews - Cochrane Central Register of Controlled Trials <September 2020>, EBM Reviews - Cochrane Database of Systematic Reviews <2005 to October 28, 2020>

Search Strategy:

--------------------------------------------------------------------------------

1	Coronavirus/ (2)

2	Coronavirus Infections/ (367)

3	(COVID-19 or COVID19).mp. (1563)

4	((coronavirus* or corona virus*) and (hubei or wuhan or beijing or shanghai)).mp. (126)

5	Wuhan virus*.mp. (2)

6	2019-nCoV.mp. (65)

7	(nCoV or n-CoV).mp. (80)

8	HCoV-19.mp. (4)

9	(SARS-CoV-2 or SARS-CoV2 or SARSCoV-2 or SARSCoV2).mp. (676)

10	(novel coronavirus* or novel corona virus*).mp. (146)

11	((coronavirus* or corona virus*) adj2 "2019").mp. (402)

12	((coronavirus* or corona virus*) adj2 "19").mp. (95)

13	(coronavirus 2 or corona virus 2).mp. (190)

14	(coronavirus* or corona virus*).ti. (201)

15	or/1-14 [COVID 19] (1687)

16	Developmental Disabilities/ (629)

17	(development* adj2 (abnormal* or atypical* or a-typical* or deficit* or delay* or deviation* or disabil* or disabled or disorder* or disturb* or handicap* or impair*)).ti,ab,kw. (4043)

18	(global adj2 delay*).ti,ab,kw. (41)

19	exp Child Development Disorders, Pervasive/ (1241)

20	(autis* or asperger*).ti,ab,kw. (3785)

21	Attention Deficit Disorder with Hyperactivity/ (2753)

22	attention deficit*.ti,ab,kw. (5292)

23	ADHD.ti,ab,kw. (4810)

24	hyperkinetic syndrome*.ti,ab,kw. (37)

25	(kanner* adj syndrome?).ti,ab,kw. (0)

26	exp Learning Disabilities/ (324)

27	((learning or scholastic*) adj3 (atypical* or a-typical* or deficit* or delay* or disabil* or disabled or disorder* or disturb* or dysfunction* or handicap* or impair* or retard*)).ti,ab,kw. (1880)

28	(alexia* or alexic* or dyslexia* or dyslexic* or reading disab* or word blindness or (verbal* adj1 agnosi*)).ti,ab,kw. (413)

29	exp Language Development Disorders/ (206)

30	((language or auditor* or semantic* or speech or speak* or talk* or verbal*) adj3 (atypical* or a-typical* or deficit* or delay* or disabil* or disabled or disorder* or disturb* or dysfunction* or handicap* or impair* or retard*)).ti,ab,kw. (3142)

31	Cerebral Palsy/ (1370)

32	((cerebral adj1 palsy) or (diplegia* adj1 spastic) or (little adj1 disease)).ti,ab,kw. (3703)

33	((brain? or central*) adj1 palsy).ti,ab,kw. (21)

34	((brain or central* or cerebral*) adj1 (paralys* or pares#s)).ti,ab,kw. (43)

35	encephalopathia infantilis.ti,ab,kw. (0)

36	exp Malformations of Cortical Development/ (66)

37	Neural Tube Defects/ (90)

38	(neural tube? adj2 defect*).ti,ab,kw. (267)

39	(acrania? or craniorachischis* or diastematomyelia* or exencephal* or iniencephal* or neurenteric cyst? or neuroenteric cyst? or spinal dysraphism* or spinal cord myelodysplasia* or (tethered adj2 cord syndrome*)).ti,ab,kw. (117)

40	spina? bifida*.ti,ab,kw. (170)

41	(cleft spine? or open spine? or rachischis* or schistorrhach* or spina? dysraphia* or status dysraphicus).ti,ab,kw. (5)

42	Intellectual Disability/ (739)

43	(intellectual* adj2 (atypical* or a-typical* or deficit* or delay* or deviation* or disabil* or disabled or disorder* or disturb* or dysfunction* or handicap* or impair* or retard*)).ti,ab,kw. (1253)

44	(brain* adj2 (atypical* or a-typical* or deficit* or delay* or deviation* or disabil* or disabled or disorder* or disturb* or dysfunction* or handicap* or impair* or retard*)).ti,ab,kw. (2057)

45	(cognitiv* adj2 (atypical* or a-typical* or deficit* or delay* or deviation* or disabil* or disabled or disorder* or disturb* or dysfunction* or handicap* or impair* or retard*)).ti,ab,kw. (16062)

46	(cognition* adj2 (atypical* or a-typical* or deficit* or delay* or deviation* or disabil* or disabled or disorder* or disturb* or dysfunction* or handicap* or impair* or retard*)).ti,ab,kw. (1181)

47	(mental* adj2 (atypical* or a-typical* or deficit* or delay* or deviation* or disabil* or disabled or disorder* or disturb* or dysfunction* or handicap* or impair* or retard*)).ti,ab,kw. (10762)

48	Down Syndrome/ (343)

49	(Down* adj2 syndrome*).ti,ab,kw. (810)

50	(mongolism* or mongoloid*).ti,ab,kw. (6)

51	("trisomy 21" or "trisomy G1" or "trisomy (G)1" or "trisomy G-1" or "trisomy GM" or "trisomy G" or "21 trisomy" or "G1 trisomy" or "G(1) trisomy" or "G-1 trisomy" or "GM trisomy" or "G trisomy").ti,ab,kw. (118)

52	"Chromosomes, Human, Pair 21"/ (26)

53	(translocat* adj1 DS).ti,ab,kw. (0)

54	Fetal Alcohol Spectrum Disorders/ (75)

55	((f?etal or f?etus*) adj2 alcohol syndrome*).ti,ab,kw. (77)

56	FASD.ti,ab,kw. (87)

57	(alcohol-related adj3 (birth defect* or birth disorder*)).ti,ab,kw. (3)

58	(alcohol-related adj3 (neurodevelopment* or neuro-development*) adj2 (deficit* or delay* or deviation* or disabil* or disabled or disorder* or disturb* or handicap* or impair*)).ti,ab,kw. (4)

59	Neonatal Abstinence Syndrome/ (90)

60	((neonat* or newborn*) adj2 (abstinen* or addiction* or withdrawal*)).ti,ab,kw. (228)

61	Fragile X Syndrome/ (90)

62	"Fragile X".ti,ab,kw. (181)

63	(FRAXA or FRAXE or "Fra(X)").ti,ab,kw. (16)

64	("Mar (X)" or "Marker X" or "Martin-Bell").ti,ab,kw. (2)

65	("X-Linked" adj2 mental retardation).ti,ab,kw. (2)

66	Angelman Syndrome/ (15)

67	(angelman* or "happy puppet" or puppet child*).ti,ab,kw. (34)

68	exp Brain Injuries/ (2151)

69	(brain* adj3 (injur* or commotio* or concuss* or damag* or trauma*)).ti,ab,kw. (8040)

70	concussion*.ti,ab,kw. (645)

71	commotio.ti,ab,kw. (10)

72	exp Stroke/ (9666)

73	stroke?.ti,ab,kw. (54988)

74	((cerebrovascular or cerebro-vascular) adj2 (accident* or arrest* or failure* or injur* or insufficienc* or insult*)).ti,ab,kw. (13438)

75	(cerebral vascular adj2 (accident* or arrest* or failure* or injur* or insufficienc* or insult*)).ti,ab,kw. (135)

76	isch?emic seizure*.ti,ab,kw. (0)

77	isch?emic cerebral attack*.ti,ab,kw. (5)

78	(apoplex* or apoplectic*).ti,ab,kw. (444)

79	((brain or cerebral) adj2 insult$2).ti,ab,kw. (81)

80	Asphyxia Neonatorum/ (209)

81	((neonat* or newborn*) adj3 (anoxi* or asphyxi* or hypoxi* or respiratory failure*)).ti,ab,kw. (937)

82	Hypoxia-Ischemia, Brain/ (208)

83	exp Infant, Premature/ (3519)

84	((preterm or pre-term or prematur* or pre-matur*) adj3 (baby or babies or infant* or neonat* or newborn*)).ti,ab,kw. (12813)

85	exp Infant, Low Birth Weight/ (2072)

86	small for gestational age?.ti,ab,kw. (963)

87	(SGA or LBW or VLBW).ti,ab,kw. (2522)

88	low* birth weight?.ti,ab,kw. (4620)

89	exp Congenital Heart Defects/ (2212)

90	((cardiac* or cardio* or heart?) adj2 (abnormalit* or anomal* or atypical* or a-typical* or defect* or deficien* or deform* or impair* or malform*)).ti,ab,kw. (2835)

91	(tetralog* adj2 fallot*).ti,ab,kw. (286)

92	((cardiac* or cardio* or heart?) adj5 (congenital* or inborn* or hereditar* or inherit*)).ti,ab,kw. (2493)

93	Intensive Care Units, Neonatal/ and (Survivors/ or Survivorship/ or "Patient Discharge"/) (25)

94	(neonat* adj2 (critical care or intensive care or ICU or ICUs) adj3 (discharg* or graduat* or surviv*)).ti,ab,kw. (105)

95	(post* adj2 (NICU or NICUs)).ti,ab,kw. (32)

96	((NICU or NICUs) adj3 (discharg* or graduat* or surviv*)).ti,ab,kw. (191)

97	or/16-96 [BRAIN/HEART DISABILITIES] (135121)

98	exp Substance-Related Disorders/ (15039)

99	(alcoholis* or alcoholic*).ti,ab,kw. (9447)

100	((binge? or binging) adj2 (alcohol* or drink*)).ti,ab,kw. (793)

101	(alcohol* adj3 (abus* or addict* or dependen* or disorder? or habituat* or misus* or mis-us*)).ti,ab,kw. (8436)

102	((narcotic* or opioid* or opiate*) adj3 (abus* or addict* or dependen* or disorder? or habituat* or misus* or mis-us* or non-medical* or nonmedical* or non-prescrib* or nonprescrib* or non-prescription* or nonprescription* or withdrawal* or (("use" or used or uses or using) adj2 (illicit* or illegal*)))).ti,ab,kw. (4722)

103	((drug? or substance?) adj3 (abus* or addict* or dependen* or disorder? or habituat* or misus* or mis-us* or "non-medical use?" or "nonmedical use?" or "non-prescribed use?" or "nonprescribed use?" or "non-prescription use?" or "nonprescription use?" or (("use" or used or uses or using) adj2 (illicit* or illegal*)))).ti,ab,kw. (19169)

104	(heroin* adj3 (abus* or addict* or dependen* or disorder? or habituat* or misus* or mis-us* or non-medical* or nonmedical* or non-prescrib* or nonprescrib* or non-prescription* or nonprescription* or withdrawal* or (("use" or used or uses or using) adj2 (illicit* or illegal*)))).ti,ab,kw. (962)

105	((hydrocodone or bekadid$2 or codinovo$2 or dico$2 or dicodid$2 or dihydrocodeinone$2 or hycodan$2 or hycon$2 or hydrocodeinonebitartrate$2 or hydrocodon$2 or hydrocon$2 or hydrocodonum$2 or robidone$2) adj3 (abus* or addict* or dependen* or disorder? or habituat* or misus* or mis-us* or non-medical* or nonmedical* or non-prescrib* or nonprescrib* or non-prescription* or nonprescription* or withdrawal* or (("use" or used or uses or using) adj2 (illicit* or illegal*)))).ti,ab,kw. (35)

106	((fentanyl or alfentanil$2 or alfenta$2 or alfentanyl$2 or beta hydroxymefentanyl or brifentanil$2 or carfentanil$2 or duragesic$2 or fanaxal$2 or fentanest$2 or fentora$2 or hypnorm$2 or limifen$2 or lofentanil$2 or mefentanyl$2 or mirfentanil$2 or ocfentanil$2 or phentanyl$2 or R-39209 or R-4263 or rapifen$2 or remifentanil$2 or sublimaze$2 or sufenta$2 or sufentanil$2 or sulfentanyl$2 or trefentanil$2) adj3 (abus* or addict* or dependen* or disorder? or habituat* or misus* or mis-us* or non-medical* or nonmedical* or non-prescrib* or nonprescrib* or non-prescription* or nonprescription* or withdrawal* or (("use" or used or uses or using) adj2 (illicit* or illegal*)))).ti,ab,kw. (111)

107	((morphine or anpec$2 or duramorph$2 or epimorph$2 or miro$2 or morfin$2 or morfine$2 or morphin$2 or morphinium$2 or morphium$2 or MS contin or morphia$2 or opso$2 or oramorph$2 or SDZ 202-250 or SDZ202-250 or skenan$2 or transmorphine$2 or trans-morphine$2) adj3 (abus* or addict* or dependen* or disorder? or habituat* or misus* or mis-us* or non-medical* or nonmedical* or non-prescrib* or nonprescrib* or non-prescription* or nonprescription* or withdrawal* or (("use" or used or uses or using) adj2 (illicit* or illegal*)))).ti,ab,kw. (442)

108	((oxycodone or bionine$2 or bionone$2 or bolodorm$2 or broncodal$2 or bucodal$2 or cafacodal$2 or cardanon$2 or codenon$2 or codix 5 or "col 003" or col003 or DETERx$2 or dihydrohydroxycodeinone or dihydrohydroxydodeinone or dihydrone$2 or dinarkon$2 or endone$2 or eubine$2 or eucodal$2 or eucodale$2 or eucodalum$2 or eudin$2 or eukdin$2 or eukodal$2 or eumorphal$2 or eurodamine$2 or eutagen$2 or hydrocodal$2 or hydroxycodeinoma$2 or ludonal$2 or m-oxy or medicodal$2 or narcobasina$2 or narcobasine$2 or narcosin$2 or nargenol$2 or narodal$2 or nsc 19043 or nucodan$2 or opton$2 or ossicodone$2 or oxanest$2 or oxaydo$2 or oxecta$2 or oxicone$2 or oxicontin$2 or oxiconum$2 or oxikon$2 or oxy ir or oxycod$2 or oxycodeinon$2 or oxycodeinonhydrochloride or oxycodone hydrochloride or oxycodonhydrochlorid or oxycodyl$2 or oxycone$2 or oxycontin$2 or oxydose$2 or oxyfast$2 or oxygesic$2 or oxyir$2 or oxykon$2 or oxynorm$2 or pancodine$2 or pavinal$2 or percolone$2 or pronarcin$2 or remoxy$2 or roxicodone$2 or roxycodone$2 or sinthiodal$2 or stupenal$2 or supeudol$2 or tebodal$2 or tekodin$2 or thecodin$2 or theocodin$2 or xtampa$2 or xtampza$2) adj3 (abus* or addict* or dependen* or disorder? or habituat* or misus* or mis-us* or non-medical* or nonmedical* or non-prescrib* or nonprescrib* or non-prescription* or nonprescription* or withdrawal* or (("use" or used or uses or using) adj2 (illicit* or illegal*)))).ti,ab,kw. (95)

109	or/98-108 [SUBSTANCE/ALCOHOL DISORDERS] (42419)

110	exp Infant/ (31489)

111	exp Child/ (55679)

112	Adolescent/ (104934)

113	(baby or babies or infant? or infanc* or neonat* or newborn* or preschool* or pre-school* or toddler?).ti,ab,kw. (73357)

114	(adolescen* or child* or pre-adolescen* or preteen* or pre-teen* or school-age* or teen or teens or teenager? or youth*).ti,ab,kw. (176301)

115	exp Pediatrics/ (675)

116	p?ediatric*.ti,ab,kw. (35594)

117	or/110-116 [INFANTS/CHILDREN/ADOLESCENTS] (304921)

118	97 and 117 [BRAIN DISABILITIES - CHILDREN/ADOLESCENTS/YOUNG ADULTS] (42949)

119	15 and 118 [COVID-19 - BRAIN DISABILITIES - CHILDREN/ADOLESCENTS/YOUNG ADULTS] (10)

120	Prenatal Exposure Delayed Effects/ (349)

121	(prenatal* or pre-natal* or preterm* or antenatal* or ante-natal* or antepartum* or ante-partum*).ti,ab,kw. (22051)

122	((before or prior) adj3 (birth* or childbirth* or child birth*)).ti,ab,kw. (408)

123	(baby or babies or infant? or infanc* or neonat* or newborn*).ti,ab,kw. (64169)

124	or/120-123 [PRENATAL EXPOSURE/NEWBORNS] (70883)

125	109 and 124 [SUBSTANCE ABUSE - PRENATAL EXPOSURE/NEWBORNS] (976)

126	15 and 125 [COVID-19 - SUBSTANCE ABUSE - PRENATAL EXPOSURE/NEWBORNS] (0)

127	119 or 126 [ALL CONDITIONS OF INTEREST] (10)

128	(2020041* or 2020042* or 2020043* or 202005* or 202006* or 202007* or 202008* or 202009* or 202010*).up. (124470)

129	127 and 128 [UPDATE PERIOD] (10)

130	129 use cctr [CENTRAL RECORDS] (9)

131	129 use coch [DSR RECORDS] (1)

***************************
