## Supplemental Data 2 for "COVID-19 in Children with Brain-Based Developmental Disabilities: A Rapid Review Update"

**Appendix 2. Grey Literature**

**List of websites consulted**

**Total number of sources**: 18
Consulted between November 5^th^ to November 13^th^, 2020
Note: Many of resources above drawn from CADTH’s Grey Matters <https://www.cadth.ca/resources/finding-evidence/grey-matters>

**Organization:** Health Canada

**Country:** Canada

**Description:** Epidemiological summary of COVID-19 cases in Canada

**Link:** <https://www.canada.ca/en/public-health/services/diseases/coronavirus-disease-covid-19.html>

**Dates consulted:** 2020-04-17, 2020-04-23, 2020-11-05

**Organization:** Canadian Agency for Drugs and Technologies in Health (CADTH)

**Country:** Canada

**Description:** Reports section

**Link:** https://www.cadth.ca/

**Date consulted:** 2020-04-17, 2020-11-05

**Organization:** Canadian Medical Association (CMA)

**Country:** Canada

**Description:** Clinical resources

**Link:** <https://www.cma.ca/cma-update-coronavirus?_ga=2.90503450.1438667280.1587060765-763235055.1585752221>

**Date consulted:** 2020-04-17, 2020-11-05

**Organization:** Health Clarity from Wolters Kluwer (global information services company)

**Country:** Multiple

**Description:** Covid-19 Resources & Tools

**Link:** <http://healthclarity.wolterskluwer.com/coronavirus-resources.html>

**Date consulted:** 2020-04-17, 2020-11-05

**Organization:** Canadian Pediatric Society

**Country:** Canada

**Description:** Covid-19 information and resources for paediatricians

**Link:** <https://www.cps.ca/en/tools-outils/covid-19-information-and-resources-for-paediatricians>

**Date consulted:** 2020-04-17, 2020-11-09

**Organization:** Evidence Aid (not-for-profit organisation)

**Country:** UK

**Description:** Covid-19 Evidence Collection

**Link:** <https://www.evidenceaid.org/coronavirus-covid-19-evidence-collection/>

**Date consulted:** 2020-04-17, 2020-11-09

**Organization:** World Health Organization

**Country:** Multiple

**Description:** Global literature on coronavirus disease

**Link:** <https://search.bvsalud.org/global-literature-on-novel-coronavirus-2019-ncov/>

**Date consulted:** 2020-04-17, 2020-11-10

**Organization:** National Institute for Health and Care Excellence (NICE)

**Country:** UK

**Description:** Coronavirus (COVID-19) Rapid guidelines and evidence summaries

**Link:** <https://www.nice.org.uk/covid-19> and <https://www.nice.org.uk/guidance/published>

**Date consulted:** 2020-04-22, 2020-11-10

**Organization:** National Institute for Health Research (NIHR)

**Country:** UK

**Description:** Innovation Observatory

**Link:** <http://www.io.nihr.ac.uk/report/>

**Date consulted:** 2020-04-22, 2020-11-11

**Organization:** National Health Service (NHS) England

**Country:** England

**Description:** Coronavirus guidance for clinicians and NHS manager

**Link:** <https://www.england.nhs.uk/coronavirus/>

**Date consulted:** 2020-04-22, 2020-11-11

**Organization:** International Resource for Infection Control (NRIC)

**Country:** UK

**Description:** Resources section

**Link:** <https://www.nric.org.uk/resources>

**Date consulted:** 2020-04-22, 2020-11-11

**Organization:** Centre for Evidence-Based Medicine (CEBM)

**Country:** UK

**Description:** Oxford COVID-19 Evidence Service

**Link:** <https://www.cebm.net/covid-19-evidence-service/>

**Date consulted:** 2020-04-22, 2020-11-11

**Organization:** Centers for Disease Control and Prevention (CDC)

**Country:** US

**Description:** COVID-19 Guidance Documents and MMWR report on Covid-19 in children

**Link:** <https://www.cdc.gov/coronavirus/2019-ncov/communication/guidance-list.html> and <https://www.cdc.gov/mmwr/volumes/69/wr/mm6914e4.htm> and <https://www.cdc.gov/coronavirus/2019-ncov/need-extra-precautions/groups-at-higher-risk.html>

**Date consulted:** 2020-04-22, 2020-04-24, 2020-04-25, 2020-11-12

**Organization:** Publons (commercial website for academics)

**Country:** Multiple

**Description:** COVID019 related publications

**Link:** <https://publons.com/publon/covid-19/?sort_by=date>

**Date consulted:** 2020-04-22, 2020-11-13

**Organization:** China Centers for Disease Control and Prevention (CCDC)

**Country:** China

**Description:** The Epidemiological Characteristics of an Outbreak of 2019 Novel Coronavirus Diseases (COVID-19) in China, 2020

**Link:** <http://weekly.chinacdc.cn/en/article/id/e53946e2-c6c4-41e9-9a9b-fea8db1a8f51>

**Date consulted:** 2020-04-22, 2020-04-25, 2020-11-13

**Organization:** Robert Koch Institute

**Country:** Germany

**Description:** Covid-19 Daily Situation Report

**Link:** <https://www.rki.de/DE/Content/InfAZ/N/Neuartiges_Coronavirus/Situationsberichte/Gesamt.html>

**Date consulted:** 2020-04-24, 2020-04-25, 2020-11-13

**Organization:** Korea Centers for Disease Control and Prevention (KCDC)

**Country:** Republic of Korea

**Description:** Press Release section

**Link:** <https://www.cdc.go.kr/board/board.es?mid=a30402000000&bid=0030>

**Date consulted:** 2020-04-22, 2020-11-13

**Organization:** Google and Goggle Scholar Search Engines

**Country:** Multiple

**Description:** -

**Link:** -

**Date consulted:** 2020-04-22, 2020-11-13

**Ongoing systematic reviews (source: PROSPERO)**

| **Author** | **Title** |
| --- | --- |
| Md Asiful Islam | Prevalence of symptoms and comorbid conditions in novel coronavirus (COVID-19)-infected adult and paediatric patients: a systematic review and meta-analysis of first three-month data of the outbreak. |
| Zeyong Yang | A systematic review and meta-analysis of the CNS comorbidities, neurotoxicity in the young, intervention treatment and clinical evolutions of patients with coronavirus disease 2019 (COVID-19) |
| Prateek Kumar Panda | Neurological complications in children with COVID-19. |

**Ongoing clinical trials (source: WHO-ICTRP)**

| **Author** | **Title** |
| --- | --- |
| Lydia A Spurr | Perceptions of the Clinical and Psychosocial Impact of Covid-19 in Patients With Neuromuscular and Neurological Disorders |
| Dr S Ramakrishnan | COVID-19 and Children with congenital heart disease |
| Jason G. Newland, MD | Supporting the Health and Well-being of Children With Intellectual and Developmental Disability During COVID-19 Pandemic |

**Preprint articles (sources: MedRxiv, BioRxiv & SSRN)**

| **Author** | **Title** |
| --- | --- |
| Luise Marino | No official help is available - experience of parents and children with congenital heart disease during COVID-19 |
| Anke Hüls | An international survey on the impact of COVID-19 in individuals with Down syndrome |
| Stankovic, Miodrag | The Serbian Experience of Challenges of Parenting Children with Autism Spectrum Disorders During the COVID-19 Pandemic and the State of Emergency with the Police Lockdown |
