## Supplemental Data 3 for "COVID-19 in Children with Brain-Based Developmental Disabilities: A Rapid Review Update"

**Appendix 3. Included Medical Conditions**

1. Attention deficit disorder; attention deficit hyperactivity disorder
2. Autism; Autistic spectrum disorder
3. Cerebral palsy
4. Childhood/developmental disability; neurodevelopmental disability
5. Congenital heart defect/disease
6. Developmental delay; global delay
7. Fetal alcohol spectrum disorder
8. Intellectual disability; Down syndrome; trisomy 21
9. Learning disability
10. Neonatal abstinence syndrome (opioid)
11. Neonatal encephalopathy
12. Neonatal intensive care unit survivors / graduates
13. Perinatal/neonatal asphyxia
14. Preterm, premature
15. Small for gestational age
16. Specific language impairment
