## Supplemental Data 4 for "COVID-19 in Children with Brain-Based Developmental Disabilities: A Rapid Review Update"

**Appendix 4. Quality Assessment**

**Table 1. Quality Assessment of case reports**

| **CASE REPORTS (n=13)** | Abasse | Bezerra | Cook | Danley | Elbehery | Gupta | Kohli | Needleman | Olfe | Piersigilli | Rodriguez | Sisman | Sumarni |
| --- | --- | --- | --- | --- | --- | --- | --- | --- | --- | --- | --- | --- | --- |
| 1. Were patient’s demographic characteristics clearly described? | Y | Y | Y | Y | Y | Y | Y | Y | Y | Y | Y | Y | Y |
| 2. Was the patient’s history clearly described and presented as a timeline? | Y | Y | Y | Y | Y | Y | Y | Y | Y | Y | Y | Y | Y |
| 3. Was the current clinical condition of the patient on presentation clearly described? | Y | Y | Y | Y | Y | Y | Y | Y | Y | Y | Y | Y | Y |
| 4. Were diagnostic tests or assessment methods and the results clearly described? | Y | Y | Y | Y | Y | Y | N | Y | N | Y | N | Y | Y |
| 5. Was the intervention(s) or treatment procedure(s) clearly described? | Y | Y | Y | Y | Y | Y | Y | Y | N | Y | Y | Y | Y |
| 6. Was the post-intervention clinical condition clearly described? | Y | Y | Y | Y | Y | N | Y | Y | Y | Y | Y | Y | Y |
| 7. Were adverse events (harms) or unanticipated events identified and described? | Y | Y | Y | Y | Y | Y | Y | Y | Y | Y | Y | Y | Y |
| 8. Does the case report provide takeaway lessons? | Y | Y | Y | Y | Y | Y | Y | Y | Y | Y | Y | Y | Y |
| **MEAN: 95%___** | **100%** | **100%** | **100%** | **100%** | **100%** | **88%** | **88%** | **100%** | **75%** | **100%** | **88%** | **100%** | **100%** |

**Legend:** Y = Yes; N = No

**Table 2. Quality Assessment of case series**

| **CASE SERIES (n=10)** | Eghbali | Krishnan | Newman | Sagheb | Schwartz | Simpson | Zhu | Paret | Xia | Zheng |
| --- | --- | --- | --- | --- | --- | --- | --- | --- | --- | --- |
| 1. Were there clear criteria for inclusion in the case series? | Y | Y | Y | Y | Y | Y | Y | Y | Y | Y |
| 2. Was the condition measured in a standard, reliable way for all participants? | Y | Y | Y | Y | Y | Y | Y | Y | Y | Y |
| 3. Were valid methods used for identification of the condition for all participants? | Y | Y | Y | Y | Y | Y | Y | Y | Y | Y |
| 4. Did the case series have consecutive inclusion of participants? | ? | ? | ? | ? | N | Y | ? | Y | ? | Y |
| 5. Did the case series have complete inclusion of participants? | ? | ? | ? | ? | N | Y | ? | Y | ? | Y |
| 6. Was there clear reporting of the demographics of the participants in the study? | Y | Y | Y | Y | Y | Y | Y | Y | Y | Y |
| 7. Was there clear reporting of clinical information of the participants? | Y | Y | Y | Y | Y | Y | Y | Y | Y | Y |
| 8. Were the outcomes or follow up results of cases clearly reported? | Y | Y | Y | Y | Y | Y | Y | Y | N | Y |
| 9. Was there clear reporting of the presenting site(s)/clinic(s) demographic information? | Y | Y | Y | Y | Y | Y | Y | Y | Y | Y |
| 10. Was statistical analysis appropriate? | NA | NA | NA | NA | NA | NA | NA | NA | NA | NA |
| **MEAN: 79%___** | **78%** | **78%** | **78%** | **78%** | **78%** | **100%** | **78%** | **78%** | **67%** | **78%** |

**Legend:** Y = Yes; N = No; ? = Unclear; NA = Not Applicable

**Table 3. Quality Assessment of cohort studies**

| **COHORT STUDIES (n=6)** | Blumfield | Desai | Götzinger | Kanburoglu | Turk | Zeng |
| --- | --- | --- | --- | --- | --- | --- |
| 1. Were the two groups similar and recruited from the same population? | Y | Y | Y | Y | Y | Y |
| 2. Were the exposures measured similarly to assign people to both exposed and unexposed groups? | Y | Y | Y | Y | Y | Y |
| 3. Was the exposure measured in a valid and reliable way? | Y | Y | Y | Y | Y | Y |
| 4. Were confounding factors identified? | NA | NA | NA | NA | NA | NA |
| 5. Were strategies to deal with confounding factors stated? | NA | NA | NA | NA | NA | NA |
| 6. Were the groups/participants free of the outcome at the moment of exposure? | Y | Y | Y | Y | Y | Y |
| 7. Were the outcomes measured in a valid and reliable way? | Y | Y | Y | Y | Y | Y |
| 8. Was the follow up time reported and sufficient to be long enough for outcomes to occur? | Y | Y | Y | Y | ? | Y |
| 9. Was follow up complete, and if not, were the reasons to loss to follow up described and explored? | Y | Y | Y | N | ? | Y |
| 10. Were strategies to address incomplete follow up utilized? | NA | NA | NA | N | NA | NA |
| 11. Was appropriate statistical analysis used? | NA | ? | Y | Y | Y | NA |
| **MEAN: 90%___** | **100%** | **88%** | **100%** | **78%** | **75%** | **100%** |

**Legend:** Y = Yes; N = No; ? = Unclear; NA = Not Applicable

**MMAT tool reference:**

Hong QN, Pluye P, Fàbregues S, Bartlett G, Boardman F, Cargo M, Dagenais P, Gagnon M-P, Griffiths F, Nicolau B, O’Cathain A, Rousseau M-C, Vedel I. Mixed Methods Appraisal Tool (MMAT), version 2018. Registration of Copyright (#1148552), Canadian Intellectual Property Office, Industry Canada
